# Immunological characteristics govern the changing severity of COVID-19 during the transition to endemicity

**DOI:** 10.1101/2020.09.03.20187856

**Authors:** Jennie S Lavine, Ottar N Bjornstad, Rustom Antia

## Abstract

As prospects for eradicating CoV-2 dwindle, we are faced with the question of how the severity of CoV-2 disease may change in the years ahead. Will CoV-2 continue to be a pathogenic scourge that, like smallpox or measles, can be tamed only by ongoing vaccination, or will it join the ranks of mild endemic human coronaviruses (HCoVs)? Our analysis of immunological and epidemiological data on HCoVs shows that infection-blocking immunity wanes rapidly, but disease-reducing immunity is long-lived. We estimate the relevant parameters and incorporate them into a new epidemiological model framework which separates these different components of immunity. Our model recapitulates both the current severity of CoV-2 and the relatively benign nature of HCoVs; suggesting that once the endemic phase is reached, CoV-2 may be no more virulent than the common cold. The benign outcome at the endemic phase is contingent on the virus causing primary infections in children. We predict a very different outcome were a CoV like MERS (that causes severe disease in children) to become endemic. These results force us to re-evaluate control measures that rely on identifying and isolating symptomatic infections, and reconsider ideas regarding herd immunity and the use of immune individuals as shields to protect vulnerable groups.

## 2 Introduction

Emerging pathogens have been invading the human host niche through history (such as smallpox emerging from an ancestral animal pox virus and rinderpest evolving to measles) to kill a substantial fraction of all people born. Recent decades have seen multiple challenges that cause severe viral disease, such as SARS, MERS, Hendra, Nipha and Ebola. Fortunately, all these are examples of emerging diseases that were locally contained. When containment is not successful, as is likely for the novel betacoronavirus SARS CoV-2 (CoV-2) [1, 2], we need to understand and plan for the transition to endemicity and continued circulation, with possible changes in disease severity due to virus evolution and build-up of host immunity and resistance.

COVID-19 is an emerging infectious disease caused by CoV-2 that has a high basic reproductive number (*R*_0_) and significant transmission during the asymptomatic phase, both of which make it hard to control [3]. However, CoV-2 is not the first coronavirus identified with human-to-human transmission: there are six other coronaviruses with known human chains of transmission, and these can provide clues to future scenarios for the current pandemic. There are four human coronaviruses (HCoVs) that circulate endemically across the globe; they cause only mild symptoms and are not a significant public health burden [4]. Another two HCoV strains, SARS CoV-1 and MERS, emerged in the past few decades and had higher case fatality ratios (CFRs) than COVID-19 but were contained and never spread widely [5, 6].

We propose a model to explore the potential changes in both transmission and disease severity of emerging HCoVs through the transition to endemicity. We focus on CoV-2, and discuss how the conclusions would differ for emerging coronaviruses more akin to SARS and MERS. Our hypothesis is that all HCoVs elicit immunity with similar characteristics, and the current acute public health problem is a consequence of epidemic emergence into an immunologically naive population in which frail age-groups have no previous exposure. We use our estimates of immunological and epidemiological parameters for endemic HCoVs to develop a quantitative transmission model and incorporate the age-dependence of severity from primary infections of emerging HCoVs. Our model explicitly considers three separate measures of the efficacy of immunity which can wane at different rates. These novel features allow us new insights into potential changes in both transmission and disease severity in the coming years (Fig 1).

**Figure 1:**
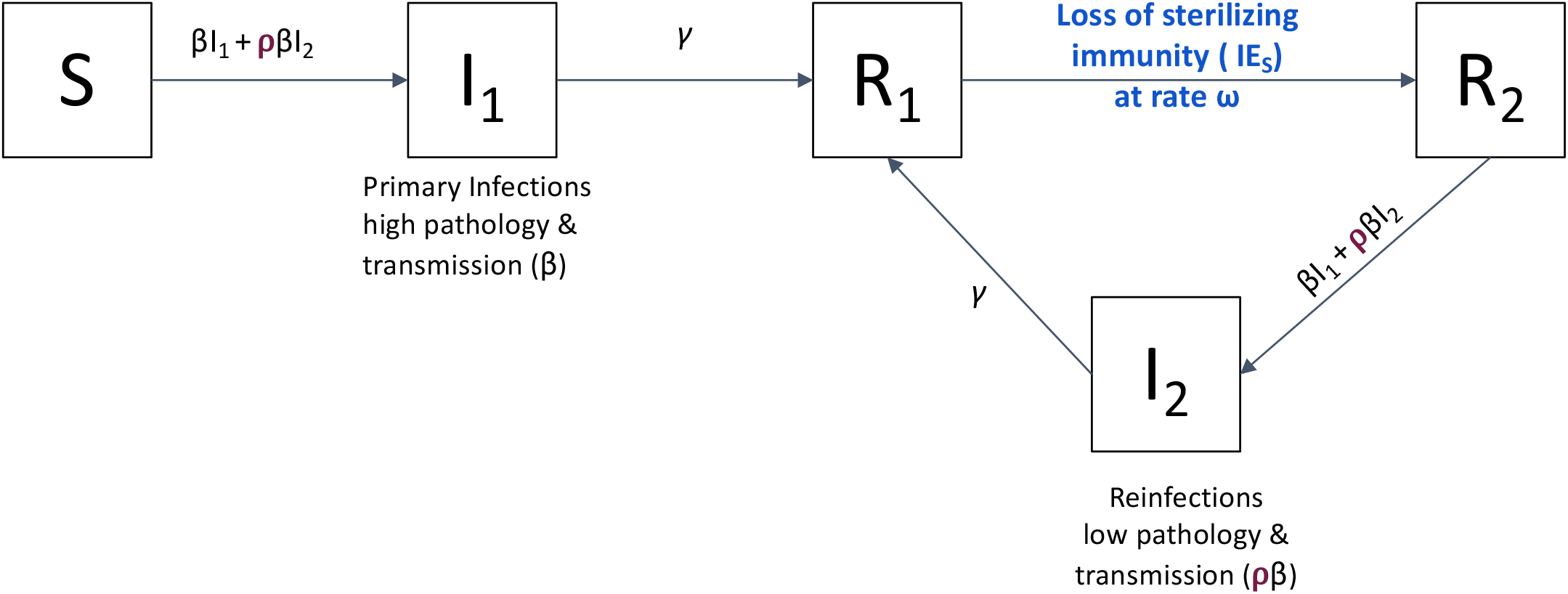
Schematic of a compartmental model for the epidemiology of CoV infections. The model incorporates the following features of natural HCoV infections: recovered individuals (*R*_1_) have sterilizing immunity (high IE_*S*_) immediately after infection, and this immunity wanes over time at rate *ω*; individuals whose sterilizing immunity has waned (*R*_2_) can become reinfected (*I*_2_), albeit with mild symptoms (IE_*P*_ is high) and reduced transmission to others (compared with transmission from primary infections, i.e., *ρ ≤* 1). We use an age-structured version of this model with parameters for demography of the US population in our simulations of transient dynamics. See SI Sec 2 for equations and details.

Building on ideas from the vaccine modeling literature, immunity may be thought to provide protection in three ways [7]. In its most robust form, “sterilizing” immunity can prevent a pathogen from replicating, thereby rendering the host refractory to reinfection. We term this property immune efficacy with respect to susceptibility, IE_*S*_. If immunity does not prevent reinfection, it may still attenuate the pathology due to reinfection (IE_*P*_) and/or reduce transmissibility or infectiousness (IE_*I*_). Experimental re-exposure studies on endemic HCoVs provide evidence that the three IE’s do not wane at the same rate [8, 9]. Callow’s experimental study [8] shows that reinfection is possible within one year (relatively short IE*_S_*); however, upon reinfection symptoms are mild (high IE*_P_*) and the virus is cleared more quickly (moderate IE*_I_*). Details the derivation of the model are in SI section 2.

## 3 Results

### 3.1 Endemic dynamics and age structure

We use available data on age-specific seroprevalence of IgM (acute response) and IgG (long-term memory) against the four circulating HCoVs to estimate parameter ranges for transmission and waning of immunity [10]. The data are plotted and further analyzed in Fig 2A. The rapid rise in seroprevelance indicates that primary infection with all four endemic HCoV strains happens early in life; and our analysis of the data gives us an estimate for the mean age of primary infection (MAPI) between 3.4 and 5.1 years, with almost everyone infected by age 15 (see SI Sec 1 for details). For most people to be infected so early in life – younger even than measles in the pre-vaccine era – there must be a very high attack rate, too high to be due only to transmission from primary infections. Steady state analysis of the model shows how this attack rate arises from a combination of high transmissibility from primary infections (i.e., high *R*_0_) and waning of sterilizing immunity and significant transmission from reinfections in older individuals. The rapid waning of sterilizing immunity is also documented in experimental HCoV infections of humans which showed that reinfection is possible one year after an earlier infection, albeit with milder symptoms (IE*_P_*) and a shorter duration (IE*_I_*) [11]. Figure 2B shows the plausible combinations of waning immunity and transmission from reinfected individuals that is required to produce the MAPI observed in Figure 2A. Table 1 shows the ranges of the parameters used in our simulations.

**Figure 2:**
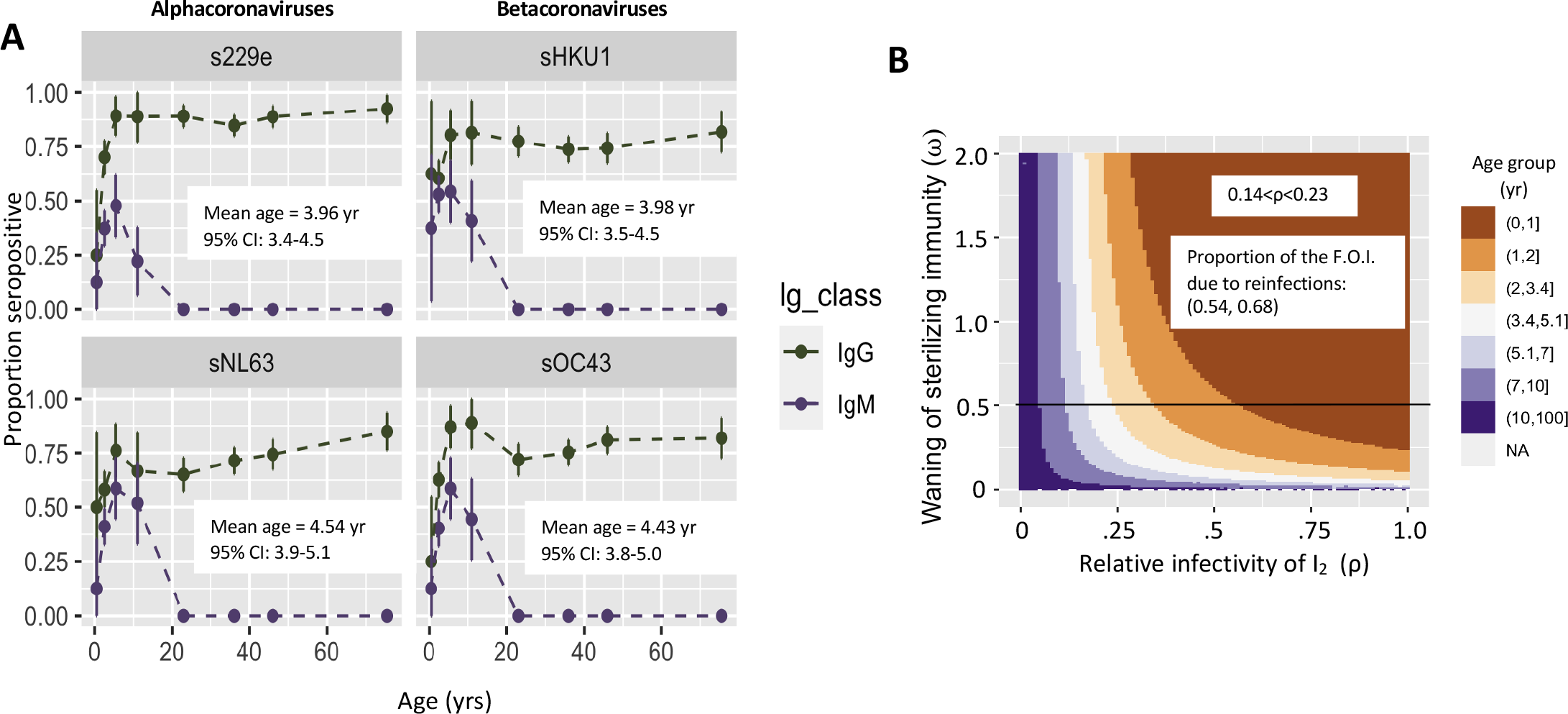
**Panel A.** Using data published by Zhou et al (2013) [10], we plotted the mean proportion seropositive for IgG (green, top lines) and IgM (purple, bottom lines) against the four endemic HCoV strains (dots connected by dashed lines) along with the 95% CIs for the proportions (vertical lines going through each dot). Additionally, we calculate the mean age of primary infection (MAPI) based on the IgM data, and show the mean and its 95% CI in text inside each panel (see SI for details). **Panel B.** We plot how the MAPI depends on the rate of waning of sterilizing immunity (*ω*, on the y-axis) and the relative transmissibility of reinfections (compared with primary infections) (*ρ*, on the x-axis). The MAPI was calculated from the equilibrium dynamics of the model shown in Fig 1 and SI Eqns 3–9 with a plausible basic reproductive number (*R*_0_ = 5) and 0 *< ω* < 2 (i.e., sterilizing immunity is as short as *≥* 0.5 yr) and 0 *< ρ* < 1 (i.e., reinfections transmit less than primary infections). See SI Sec 2.1 for details. The white band in Panel B indicates the plausible combinations of values of *ρ* and *ω* consistent with the MAPI for HCoVs estimated in Panel A. If additionally we restrict the duration of sterilizing immunity to at most two years (*ω >* 0.5, plot area above the black line in Panel B), then the plausible range for the relative transmissibility of reinfections is 0.14 *< ρ* < 0.23. Reinfected individuals cause between 54 and 68% of primary infections. (See SI Fig 1 for parallel figures calculated at the extremes of plausible values for *R*_0_ as defined in Table 1 (i.e., *R*_0_ = 2 and *R*_0_ = 10).)

**Table 1:** Characteristics of coronavirus-immune interactions and relevant parameter ranges

| Characteristic & symbol | Estimates from literature | Value (range) | Citations |
| --- | --- | --- | --- |
| Primary infectious period ($1/\gamma$), in days | $\geq 5.6$ d<br>$\sim 10$ d | 9 d | [8]<br>[12] |
| Primary transmissibility ($R_0 = \frac{\beta}{\gamma+\mu}$) | 4-9 <sup>c</sup> | 2-10 | [13] |
| Secondary transmissibility (relative to primary, $\rho$) | 0.35<br>0.04-0.97 | 0-1 | [8]<br>Fig 2 & SI |
| Duration of sterilizing immunity ($1/\omega$), in years | 0.91 yr<br>1.67 yr<br>0.5-2 yr | 0.5-10 yr | [8] & SI<br>[8] & SI<br>[11, 14] |
| Relative pathology of reinfections | mild |  | [8] |
| Age-specific CFR (primary infections) | SARS<br>MERS<br>COVID-19 | see Fig 4 | [15]<br>[16]<br>[17] |

## 3.2 Transition from pandemic to endemic dynamics

At the beginning of an outbreak, the age distribution of cases mirrors that of the population (Fig 3A). Once the demographics of infection reaches a steady state, however primary cases occur almost entirely in babies and young children, who in the case of COVID-19, experience a low CFR. Reinfections in older individuals are predicted to be common and contribute to transmission, but in this steady-state population, older individuals, who would be at risk for severe disease from a *primary* infection, have acquired disease-reducing immunity following infection during childhood.

**Figure 3:**
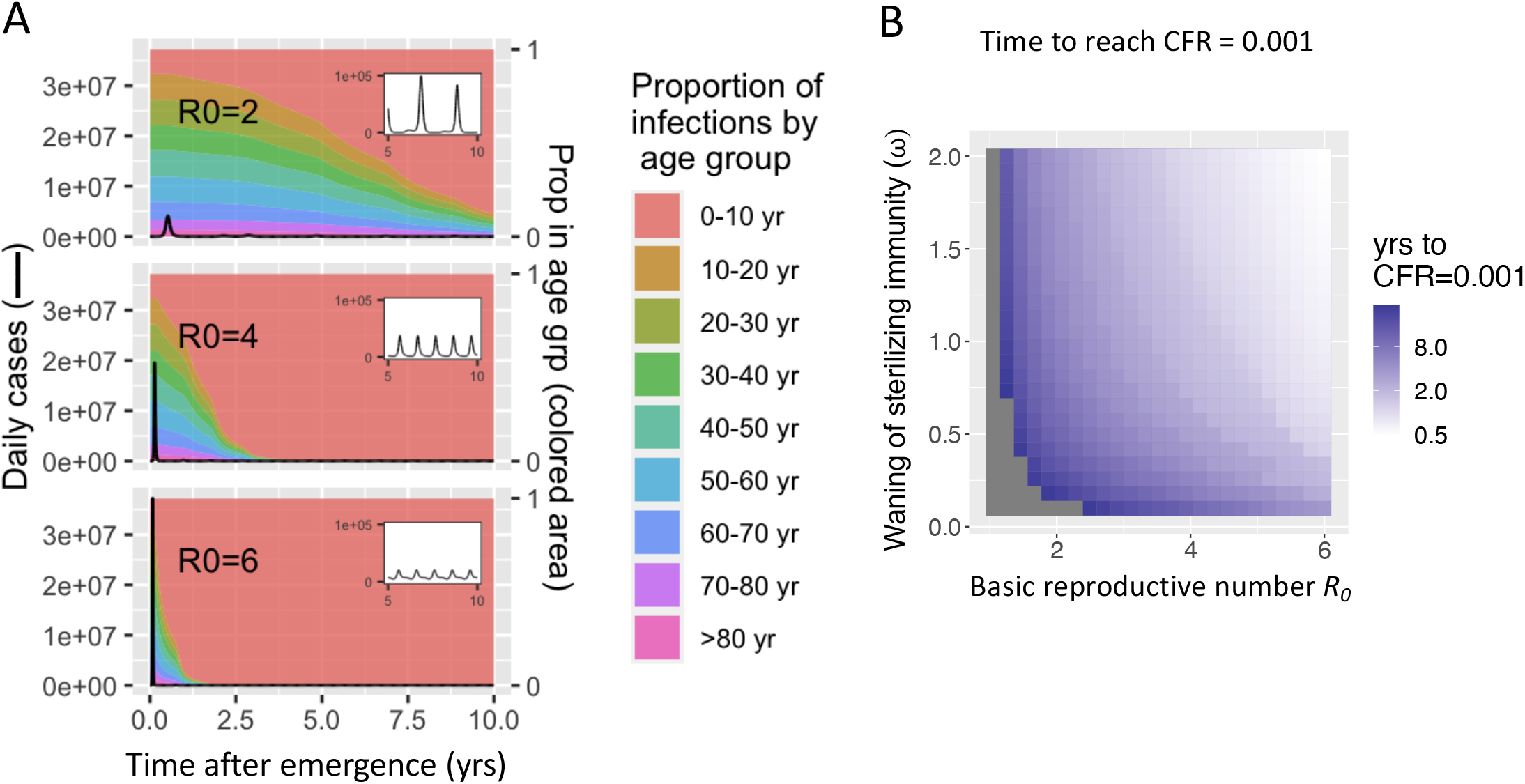
Simulations for the transition from epidemic to endemic dynamics for emerging CoVs. Simulations from an extension of the model presented in Fig 1 that includes age structure. Demographic characteristics such as the population’s age distribution, birth, and age-specific death rates are taken from the USA, and seasonality is incorporated via a sinusoidal forcing function (see SI Sec 2.2). **Panel A**. The black line representing the number of infections per day shows an initial peak and subsequent transition to endemic dynamics with much lower numbers of infections (with the endemic dynamics from years 5 to 10 shown in the inset). The three plots in A show that a higher *R*_0_ results in a larger and faster initial epidemic and more rapid transition to endemic dynamics. We also show (on the same plot) how the proportion of primary cases in different age groups changes over time (plotted in different colors), and how the transition from epidemic to endemic dynamics results in primary cases being restricted to the lower age groups. Parameters for simulations: *ω* = 1 and *ρ* = 0.7. In **Panel B**, we plot how long it takes for the average CFR (calculated as a 6 month moving average) to fall to 0.001, the CFR associated with seasonal influenza. Grey areas represent simulations where the CFR did not reach 0.001 within ten years. We see that the time taken for the CFR to decline to that of influenza decreases as the transmissibility (*R*_0_) increases and the duration of sterilizing immunity becomes shorter. Results are shown for *ρ* = 0.7. See SI Sec 2.3 and SI Figs 3–5 for sensitivity analyses and model specifications.

The overall CFR for CoV-2 is predicted to drop dramatically, eventually falling below that of seasonal influenza (approximately 0.001, see Fig 4 B top panel) due to the age-structure of primary infection. The time it takes to complete the shift in CFR en route to endemicity depends on both transmission (*R*_0_) and loss of immunity (*ω* and *ρ*) (Fig 3B and SI Fig 1).

**Figure 4:**
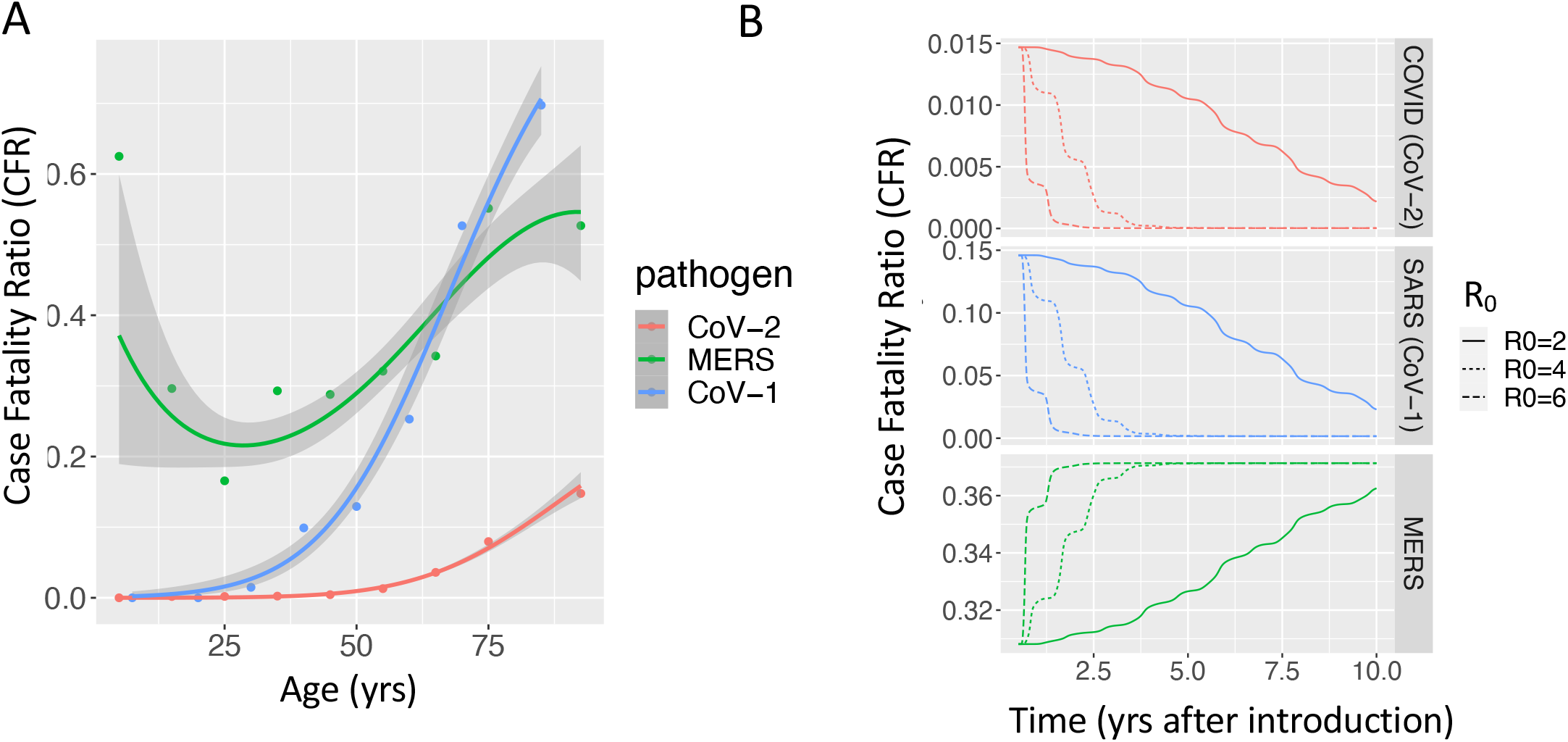
The age dependence of the CFR determines how the overall CFR changes during the transition from epidemic to endemic dynamics for emerging CoVs. **Panel A** shows the age dependence of the CFRs for the three emerging CoVs. CoV-1 and CoV-2 have a J shaped profile, with a monotonic increase in CFR with age. In contrast, the age dependence of the CFR for MERS is U shaped, with high mortality in the younger as well as older age groups. Details of the statistical smoothing are described in SI Sec 5. **Panel B** shows how the overall CFR (computed from model-predicted infections in each age group multiplied by the age-specific CFRs for the different CoVs) changes with the transition from epidemic to endemic dynamics. We see the overall CFR declines as CoV-2 transitions to endemicity, and we predict a similar trend should a CoV-1 (SARS)-like viurs spread widely. In contrast, the model predicts the CFR for a MERS-like virus would increase with time; this increase is a consequence of the shape of the age-dependence of the CFR for MERS.

In the absence of vaccination or treatment, high transmission is predicted to lead to a high case load and death rate in the initial epidemic phase during the first few years following emergence (Fig 3 and SI Figs 3 & 4). The total number of deaths is largely independent of *R*_0_ (see SI Fig 6). Slowing down the epidemic through social distancing measures that lower *R*_0_ flattens the curve, thus delaying infections and preventing the majority of deaths from happening early on, affording critical time for the development of an effective vaccine or treatment. However, as the time scales of disease spread and waning of immunity meet, transmission from reinfected individuals (likely asymptomatic or mild) to the vulnerable is predicted to increasingly drive excess deaths (SI Fig 5).

Our prediction of mild CoV-2-associated pathology in the long run stems from the low CFR for primary infections in young children. Were a more pathogenic emerging coronavirus to become endemic, for example one with an age-dependent CFR similar to CoV-1, we still expect a low disease burden in the long run (Fig 4). However, data suggest not all emerging HCoVs follow this optimistic pattern; the overall CFR of an endemic MERS-like virus would increase during the transition to endemicity because disease severity is high in children, the age group expected to experience the bulk of primary cases during the endemic phase.

## 4 Discussion

The key result from our new model framework that explicitly recognizes that functional immunity to reinfection, disease and shedding are different is that, in contrast with infections that are severe in childhood, CoV-2 could join the ranks of mild, cold-causing endemic human coronaviruses in the long run. A critical prediction is that the severity of emergent CoVs once they reach endemicity depends only on the severity of primary infection in children (Fig 4) because all available evidence suggests immunity to HCoVs has short IE_*S*_, moderate IE_*I*_ but strong IE_*P*_ such that childhood infection provides protection from pathology upon reinfection in adulthood. Strain-specific virulence factors, such as the shared cellular receptor, ACE-2, to which CoV-1, CoV-2 and the endemic strain NL63 all bind [18, 19, 20, 21], may affect the CFR during the emergence phase but have little impact on the severity of disease in the endemic phase. Because the four endemic HCoVs have been globally circulating for a long time and almost everyone is infected at a young age, we cannot ascertain how much pathology would result from a primary case of any of these in an elderly person.

The key insights come from how our model explicitly incorporates different components of immunological protection with respect to susceptibility, pathology and infectivity (IE_*S*_, IE_*P*_ and IE_*I*_) and their different rates of waning. In our analysis we hypothesized that the rates for CoV-2 are comparable to endemic HCoVs. Testing this assumption for CoV-2 will follow from ongoing and future longitudinal studies that combine clinical observations with measures of virus titers, virus shedding and antibody and T cell immunity following primary, secondary and subsequent infections of individuals. In our projections, we make the simplifying assumption that immune memory following a single infection generates protection from pathology (high IE_*P*_). However, we expect our results to be robust to a more gradual build-up of immunity in childhood when disease is mild. The details of the change in overall CFR through the transient period will be impacted by a wide array of factors, such as age-specific human contact rates [22] and susceptibility to infection [23], as well as improvement in treatment protocols, hospital capacity, and virus evolution. The qualitative result of mild disease in the endemic phase is robust to these complexities, but quantitative predictions for the transient phase will depend on a careful consideration of these realities and how they interact with the dynamics of infection and components of immunity.

The changes in the CFR over time predicted by the model have implications for vaccination strategy against current and future emerging HCoVs. The widespread (pandemic) circulation of any novel pathogenic HCoV will invariably require rapid generation and deployment of a vaccine. In the longer term, however, the necessity for continual vaccination will depend on the age-dependence of the CFR. If primary infections of children are mild (CoV-1 and CoV-2), continued vaccination may not be needed as primary cases recede to mild childhood sniffles. If, on the other hand, primary infection is severe in children (as for MERS), then vaccination of children will need to be continued.

Our model provides strategic insights into plausible transitions from pandemic to endemic dynamics for CoV-2, which have implications for control measures during a likely high-CFR transient period. These insights are outlined in Table 2. The findings presented here suggest that using symptoms as a surveillance tool to curb the virus’s spread will become more difficult, as milder reinfections increasingly contribute to chains of transmission and population level attack rates. In addition, infection or vaccination may protect against disease but not provide the type of transmission-blocking immunity that allows for shielding [24] or the generation of long-term herd immunity [2].

**Table 2:** Effects of separated IEs model on control strategies for SARS-CoV-2

|  | Effects Predicted by the Model | Possible Solutions |
| --- | --- | --- |
| Quarantine + contact tracing | Waning transmission-reducing immunity ($IE_S$ and $IE_I$) with long-lasting protection against severe disease ($IE_P$) leads to an increase in asymptomatic re-infections. Identifying and quarantining index cases becomes more difficult. Localized outbreaks are more difficult to control, particularly in places with a large first wave of infections (SI Sec 4). | Widespread / universal testing regardless of symptoms |
| Shielding & immunity passports | Given that prior infection and the presence of CoV-2-specific antibodies may indicate stronger $IE_P$ than $IE_S$ or $IE_I$, this strategy is a double-edged sword. Those with antibodies may be less likely than those without to get infected and transmit, but they may also be more likely to have an asymptomatic infection that can spread without their knowledge of it. The chance of this scenario increases with time since infection. It may not be possible to rely on seropositive individuals as caregivers to shield the elderly and vulnerable as has been proposed [24], but they are a good choice for frontline workers or those caring for COVID patients because they are less likely to have severe pathology following reinfection. | • Identify immune surrogates of $IE_S$, $IE_P$ and $IE_I$.<br>• Time stamps on immunity passports. |
| Social distancing & PPE use | Both social distancing and everyday use of personal protective equipment (PPE) reduce the spread of infection (i.e., reduce $R_0$) regardless of symptoms and therefore their efficacy does not depend on detecting symptoms. | Increase PPE use to maintain local $R_0 < 1$ with businesses as open as possible. |
| Vaccination | Vaccination, like natural infection, may not provide long-lasting transmission-blocking immunity. Consequently herd immunity will not be achieved, and the traditional strategy of vaccinating high-contact rate groups will not provide long-term protection to the rest of the population. Instead, vaccination should target those most vulnerable to severe disease (i.e., the elderly) to reduce the overall CFR during the transition to endemicity and essential workers most frequently exposed (i.e., front-line workers). | • Target vaccination toward the elderly, frontline workers, and other vulnerable populations.<br>• Generate a ‘smart vaccine’ that produces better-than-natural, transmission-blocking immunity. |

From an ecological and evolutionary perspective, our study opens the door to questions regarding the within-host and between-host dynamics of human immunity and pathogen populations in the face of IE’s with different kinetics. It also opens the question of how these IE’s interplay with strain cross-immunity, which is likely relevant within the alpha- and beta-coronaviruses. Considering data and model predictions from emergence through endemicity of HCoVs revealed a framework for understanding immunity and vaccination that may apply to a variety of infections, such as RSV and seasonal influenza, which share similar age distributions and immune responses.

## Data Availability

Analysis involves data from literature. It is available in the supplementary information

## Supplement

### 1 Serostudy analysis details

We used data from a 2013 study [1] on the seroprevalence of IgG and IgM antibodies against the Spike (S) protein of the four endemic HCoV strains in a cross-section of the population in Beijing, none of whom exhibited symptoms. A striking feature of these data is that IgM titers are undetectable in all of the more than 140 subjects ages 15 yr and older. This observation combined with general knowledge of the the IgM response [2] suggests that S-specific IgM is only elicited during primary infection. Additionally, IgM titers decay quickly, which makes IgM seropositivity a useful marker of recent primary infection.

We calculate error bars on the seroprevalence for both IgM and IgG and estimate the mean age of primary infection (MAPI) for each of the four strains from the IgM data. We assume the data for both IgM and IgG stem from a binomial process where the probability of seropositivity in age group *j* is *p_j_*, estimated by ^*p_j_* and the sample size for each age group is *N_j_*. The 95% confidence interval around the mean proportion seropositive for each age group is then

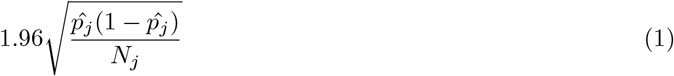

We further estimate the MAPI using only the IgM data (we assume that the cases are uniformly distributed within each age group – a more accurate estimate could be obtained from the raw data with smaller age bins; unfortunately we were unable to gain access to it). For each HCoV strain, *s*, we create a vector, *A_s_* with length *L* containing the ages ages of IgM seropositivity (using the midpoints of each age range). We assume this is a reasonable reflection of ages of first infection, as IgM titers increase only during primary infection and decay in a matter of weeks [3]. The 95% CI for the MAPI for each strain, *s*, is therefore estimated by

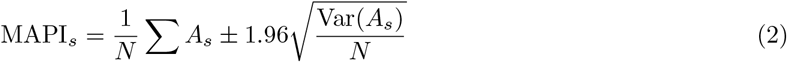

### 2 Model derivation

We derived the model presented in Fig 1 by combining three key sources of information: (1) classic SIRS disease transmission models, (2) separated functional immune efficacies (IE’s, [4]), and (3) a human reinfection experiment with HCoV 229e in fifteen healthy adult volunteers [5].

**Figure 1:**
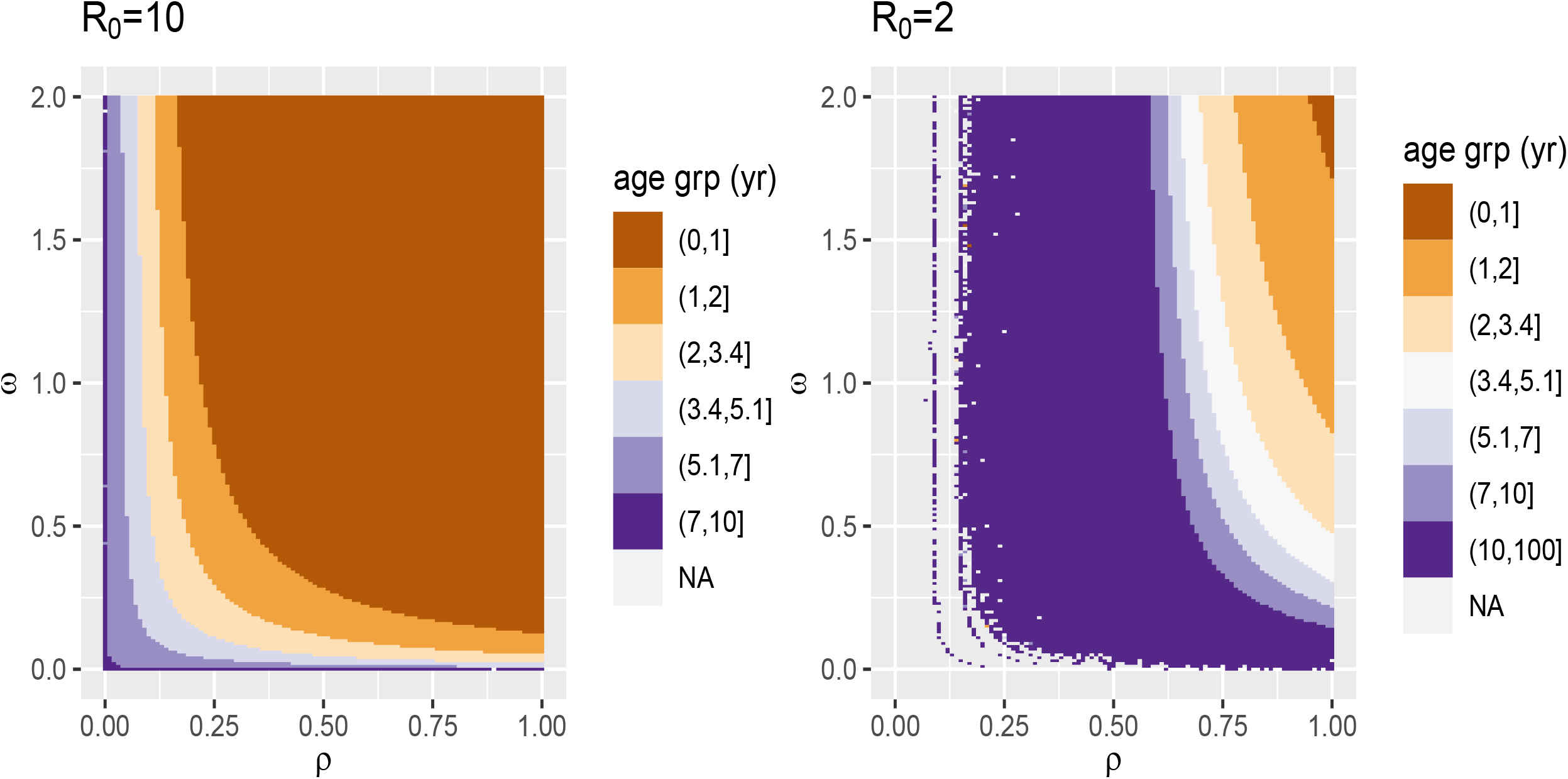
Mean age of primary infection

In aforementioned reinfection experiment, all subjects had serum specific antibodies at the start, suggesting that participants had already experienced a primary infection. The first experimental exposure resulted in viral replication and a boosting of IgG titers in ten of the fifteen participants; the group that supported viral replication had lower serum specific IgG, IgA and nasal IgA levels prior to exposure and shed virus for on average 5.6 days; eight out of ten had cold symptoms. Antibody titers increased significantly approximately a week after infection and then slowly decayed over the course of the next year. Among the five participants who did not get infected following exposure at the beginning of the trial, antibody titers remained relatively constant following exposure; however their IgG titers did drop significantly by the end of the year.

One year after the initial exposure, fourteen of the fifteen participants were re-exposed. All five who had not sustained an infection the first time became infected (their *IE*_S_ waned) and one developed cold symptoms (*IE*_P_ was still strong for four of the five). Six of the nine who had supported viral replication a year earlier sustained an infection following exposure the second time (their *IE*_S_ waned within the span of year) but none developed cold symptoms (they retained *IE*_P_); the other three did not become infected following exposure. Among all who got infected at the second time point, the mean duration of viral shedding was only 2.0 days, suggesting that *IE*_I_ had not waned completely.

Based on these observations, the following are the basic equations that correspond to Fig 1.

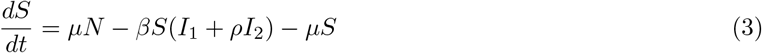

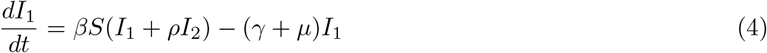

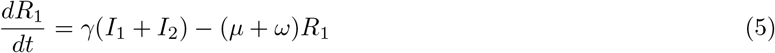

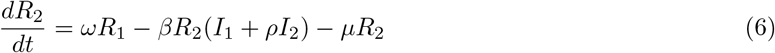

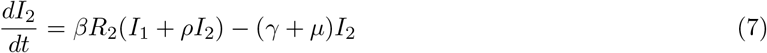

#### 2.1 Steady-state analysis

The above equations are used for steady state analysis of the predicted mean ages of primary infection (MAPIs). The model-predicted MAPI for a given set of parameters is calculated as the waiting time from birth to first infection according to the following equation:

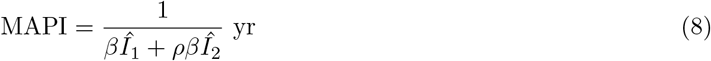

where *Î_i_* is the equilibrium proportion of the population in class *I_i_*.

The equilibrium values are calculated by first running a short (100 iterations = 1/10 yr) numerical simulation using a wrapper for the R function lsoda and then using the final values as estimates to start the Newton-Raphson method to find equilibria as implemented in the R package rootSolve, function stode.

We additionally calculate the proportion of cases caused by reinfections as follows:

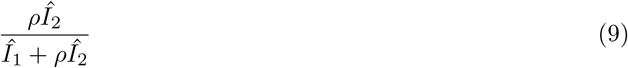

In addition to the results shown in the main text for *R*_0_ = 5, we here show figures parallel to Fig 3b for *R*_0_ = 2 and *R*_0_ = 10 (SI Fig 1).

**Figure 2:**
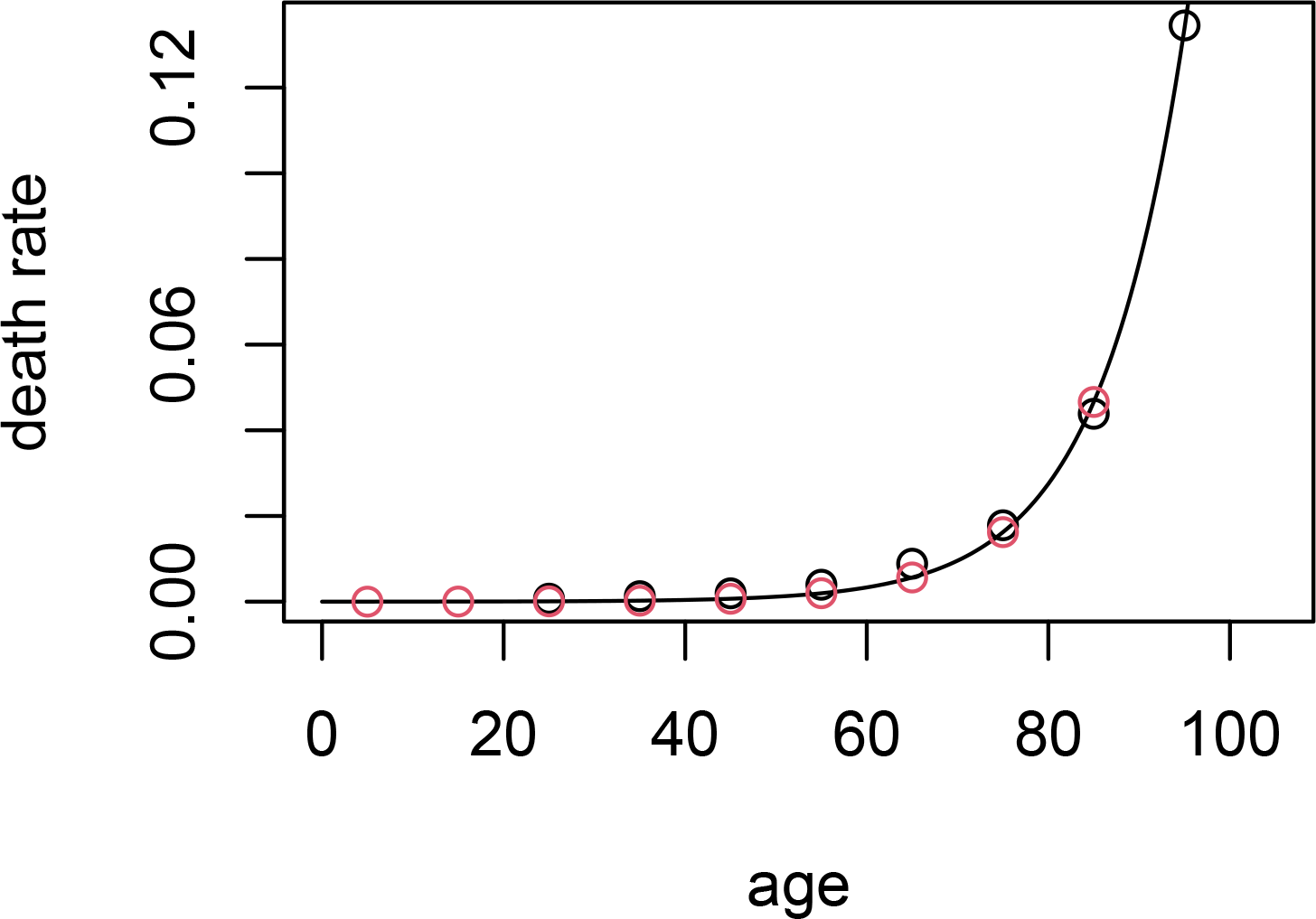
Age specific death rate in the US

**Figure 3:**
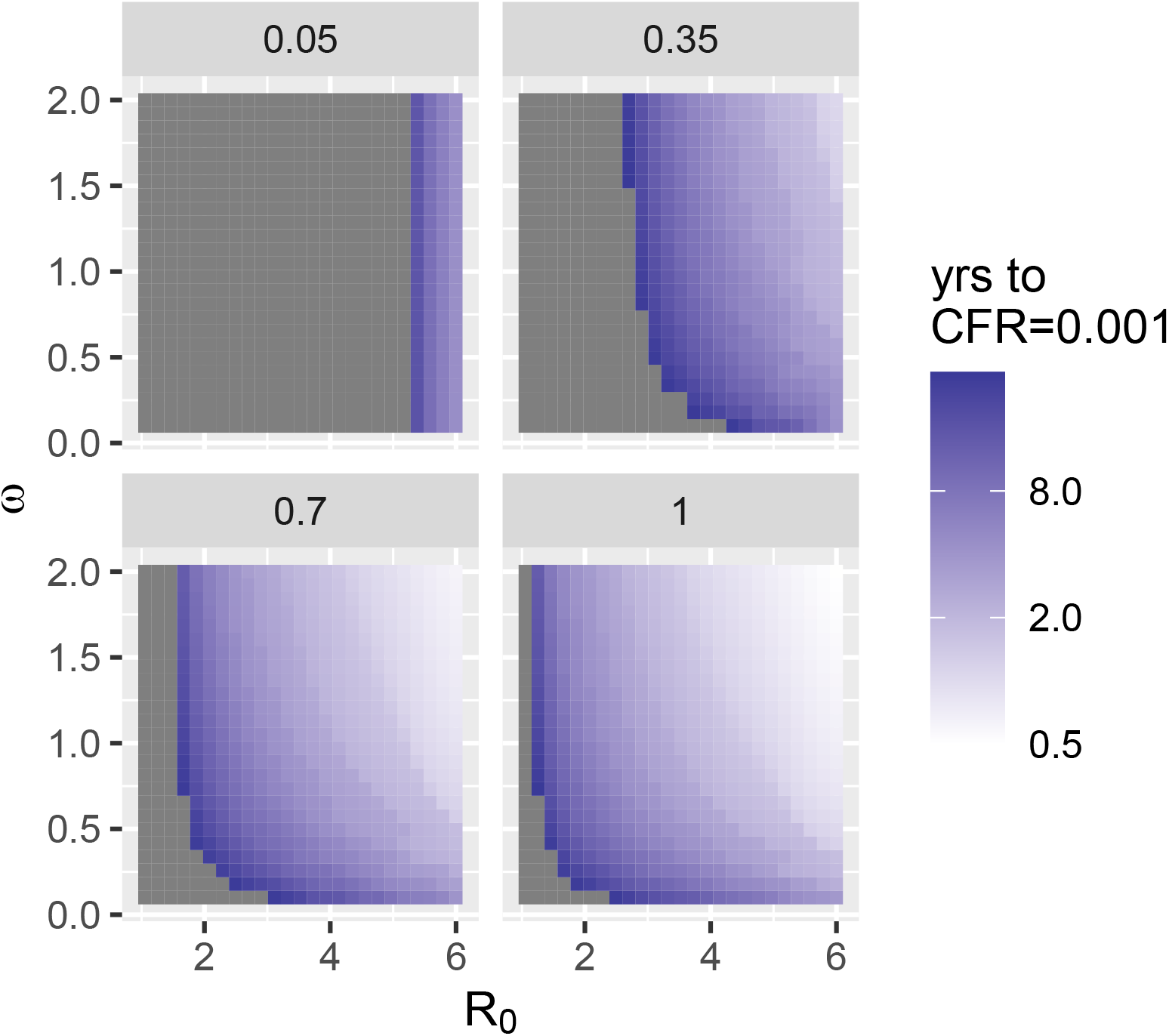
Time to CFR = 0.001, parallel to main text Fig 3b

#### 2.2 Transient to endemic dynamic simulations

To incorporate seasonality with a peak in early January (modeling on influenza and seasonal coronaviruses) and the introduction of the virus in early March (as was approximately observed with CoV-2 in the US), cases are introduced at t = 0 and we allow *β* to fluctuate annually according to

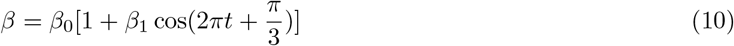

*β*_0_ is the mean value of *β*, and *β*_1_ is the amplitude of the sin wave. All results shown here use *β*_1_ = 0.2 (SI Figs 3–6 and Fig 3 in the main text).

Additionally, to incorporate the age-specific case fatality rates, death rates (*δ*), and the current age distribution of the US population, we separate each immune state, *X*, into nine age classes *X_j_* with 10-yr widths for the first eight. The aging rates correspond to the width of the age classes; the death rates and age distribution are taken from US data. This yields the following equations:

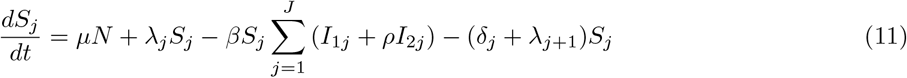

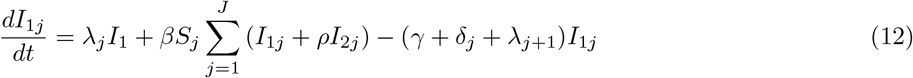

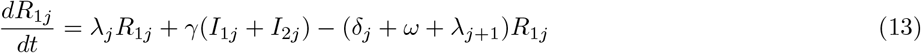

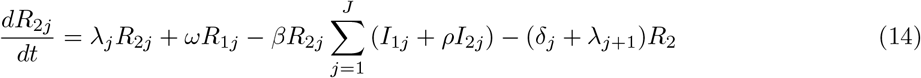

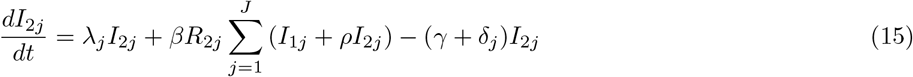

where 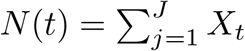 is the total population size at time *t*. The birth rate, *µ*, is a vector of length nine, containing the overall population birthrate (based on demographic data) followed by zeros for all subsequent age classes (i.e., people are only born into the youngest age class). The age-specific death rates, *δ_j_*, are fixed at values estimated from demographic data. The aging rates, *λ_j_*, are contained in a vector of length ten: (0, 0.1, 0.1, 0.1, 0.1, 0.1, 0.1, 0.1, 0.04, 0). We fix *γ* at 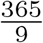 corresponding to an infectious period that lasts on average nine days. *β*_0_ is calculated according to 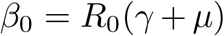. We consider a range of values of *R*_0_ (2-10), *ω* (0-2), and *ρ* (0-1).

The age-specific death rates were inferred from CDC data Health Statistics [6], and were calculated as follows:

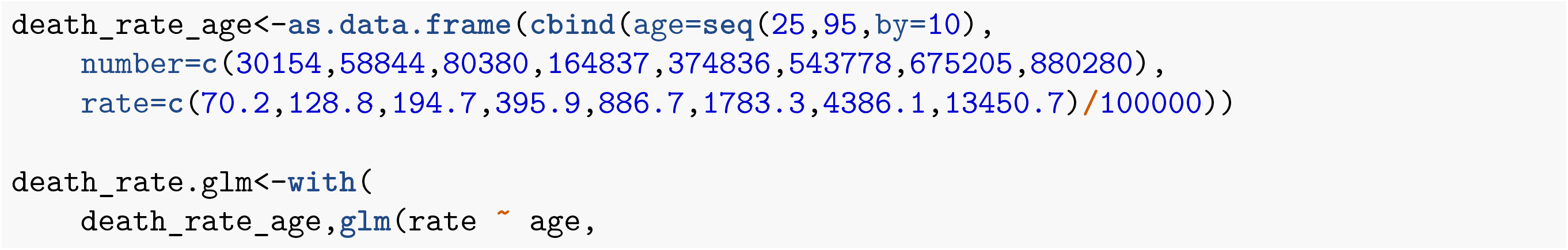
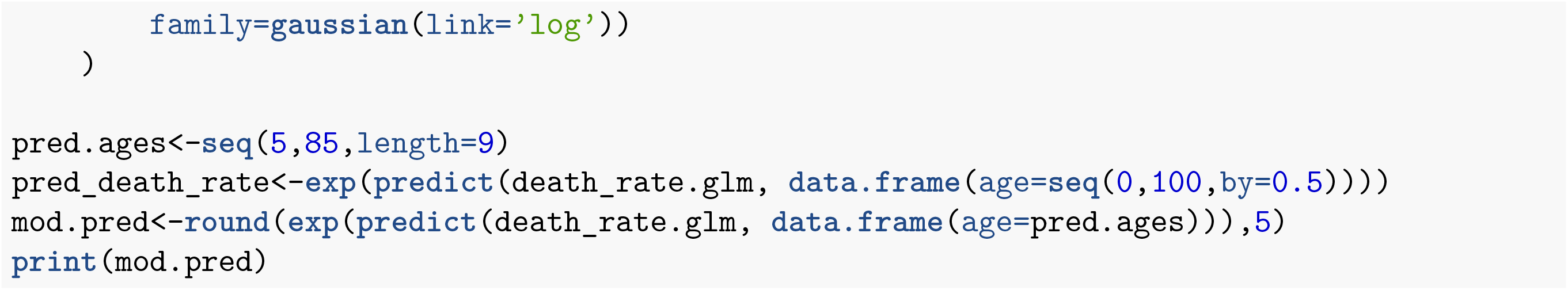

The ages used in the model prediction for death rates were the midpoints of the age classes (e.g. 5, 15, 25yr, etc). We used the death rate for 85 yr olds as the highest death rate.

The initial conditions were set so the population was distributed into the nine susceptible classes according to the age distribution of the US population [7].

One infected individuals was seeded into each age group’s *I*_1_ class.

#### 2.3 Calculating infections and the case fatality rate from simulations

The following steps were used to calculate the number of daily infections and 6-month moving average case fatality rate (CFR):

1. Numerically integrate equations with chosen parameters and initial conditions as described above using the R function lsoda with a time step of one day (1/365 yr).
2. Calculate the probability of staying in *I*_1_ for a timestep of one day given that you’re already there. In the simplest version of the model we consider here, this can be calculated using the cumulative distribution function, *F*:

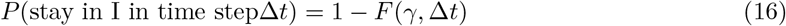

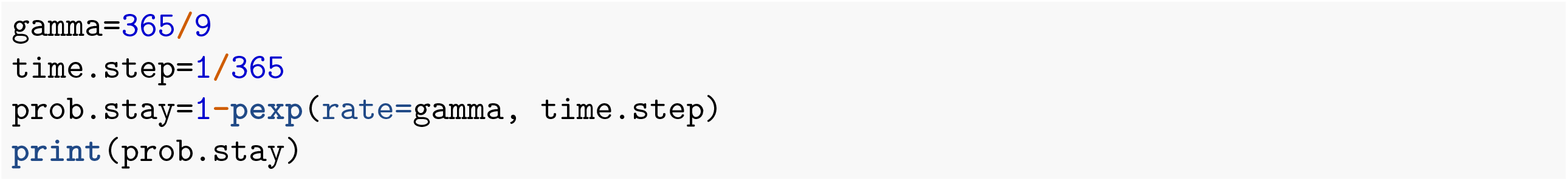 ## [1] 0.8948393
3. For each age group, calculate the number of new primary infections in each time step, *X_t_*

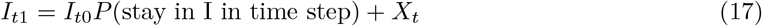

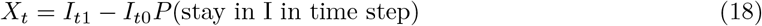

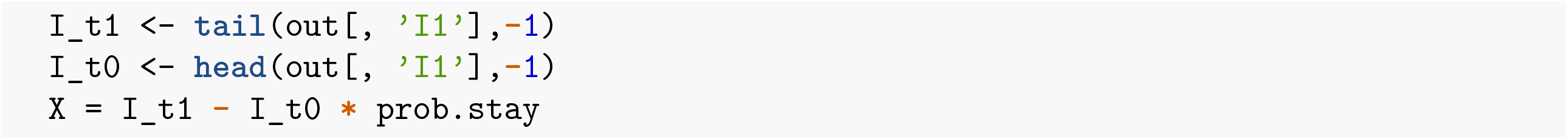
4. Calculate the projected number of HCoV-induced deaths in each age group based on the age-specific CFR’s.

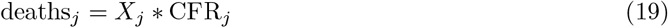
5. For every 6-month window, calculate the overall CFR beginning six months into the pandemic

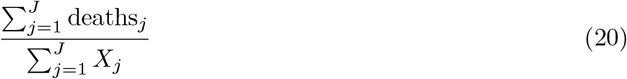

The key result that the overall CFR drops to something akin to seasonal influenza (0.001), is robust across a wide range of values for *R*_0_, *ω* and *ρ* for COVID-like CFRs.

We additionally show results for simulations with different initial conditions, in which 2000 infections were seeded into each age groups approximating the situation when the pandemic was under control in the U.S. (i.e., there had been already been many infections and *R*_0_ was close to 1). The CFR evolves very similarly to the case where only nine cases are introduced.

Additionally, we see that keeping *R*_0_ below a threshold value (in these simulations approximately 2, e.g., by social distancing and the use of Personal Protective Equipment) allows us to stop the majority of deaths from happening early on, buying time for the development of an effective vaccine and/or treatment (SI Fig 5).

**Figure 4:**
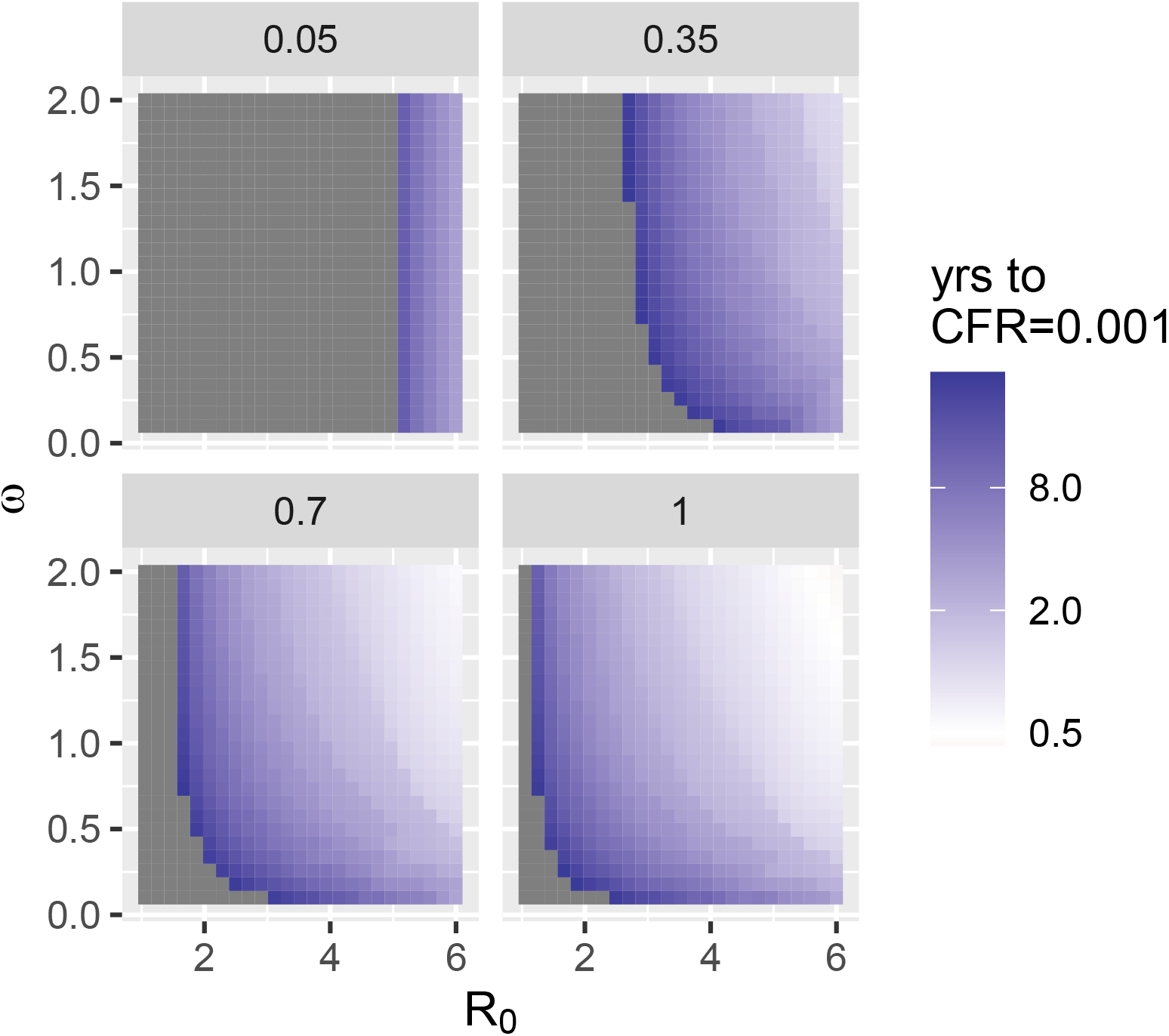
Effect of initial outbreak on time to CFR = 0.001

**Figure 5:**
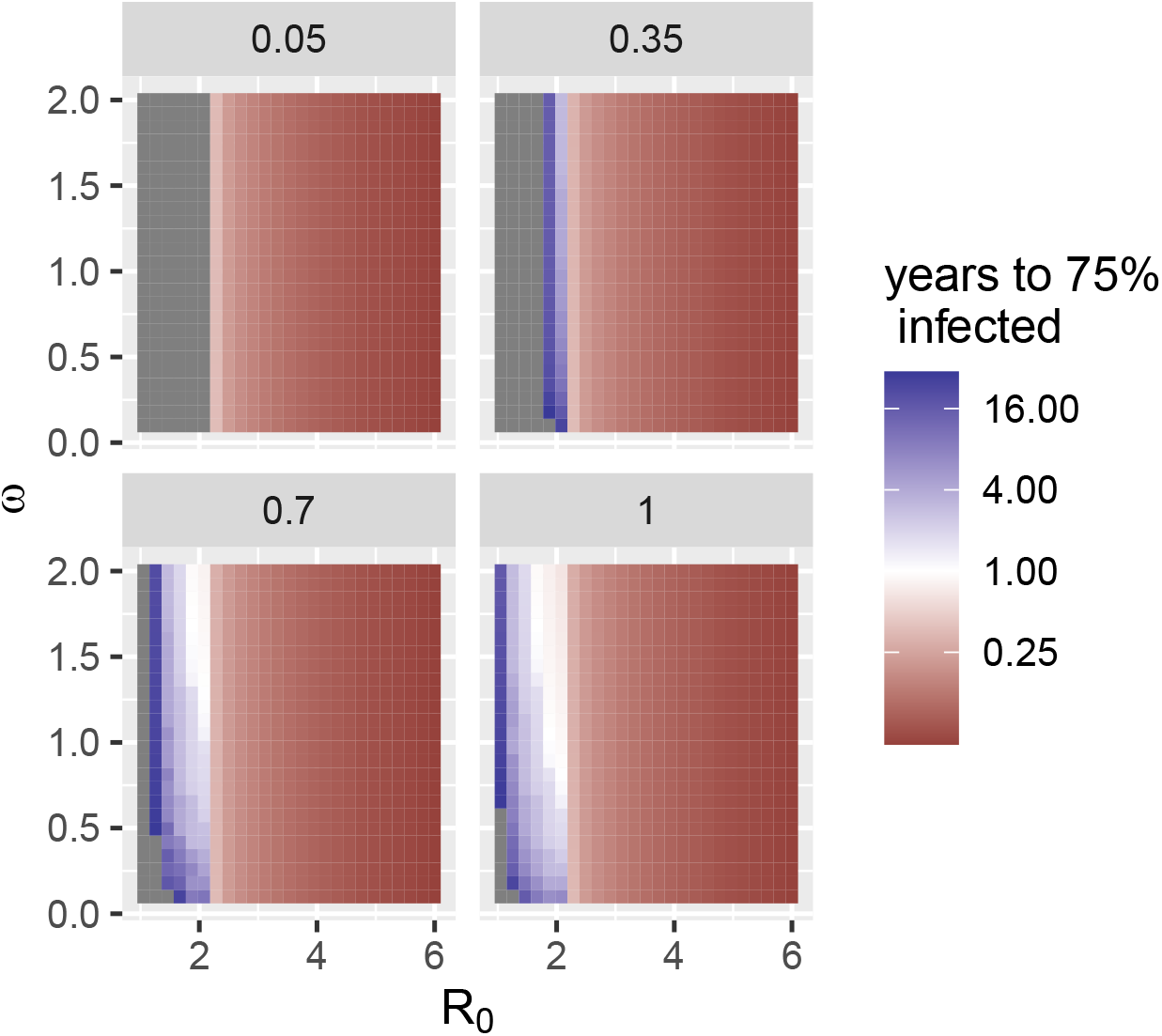
The time it takes (in yr) for 75% of the initial population size to become infected. For *R*_0_ *>* 2 the vast majority of infections occur in the first year. By slowing the epidemic down (i.e., decreasing *R*_0_, infections are delayed buying time for the development of a vaccine and/or treatment.

However, as we slow down the epidemic by decreasing *R*_0_, the time scales of disease spread and immune waning meet, and reinfections increasingly drive excess deaths, and the total number of infection-induced deaths within the first decade after emergence is very similar regardless of how fast the infection is spreading (SI Fig 6).

**Figure 6:**
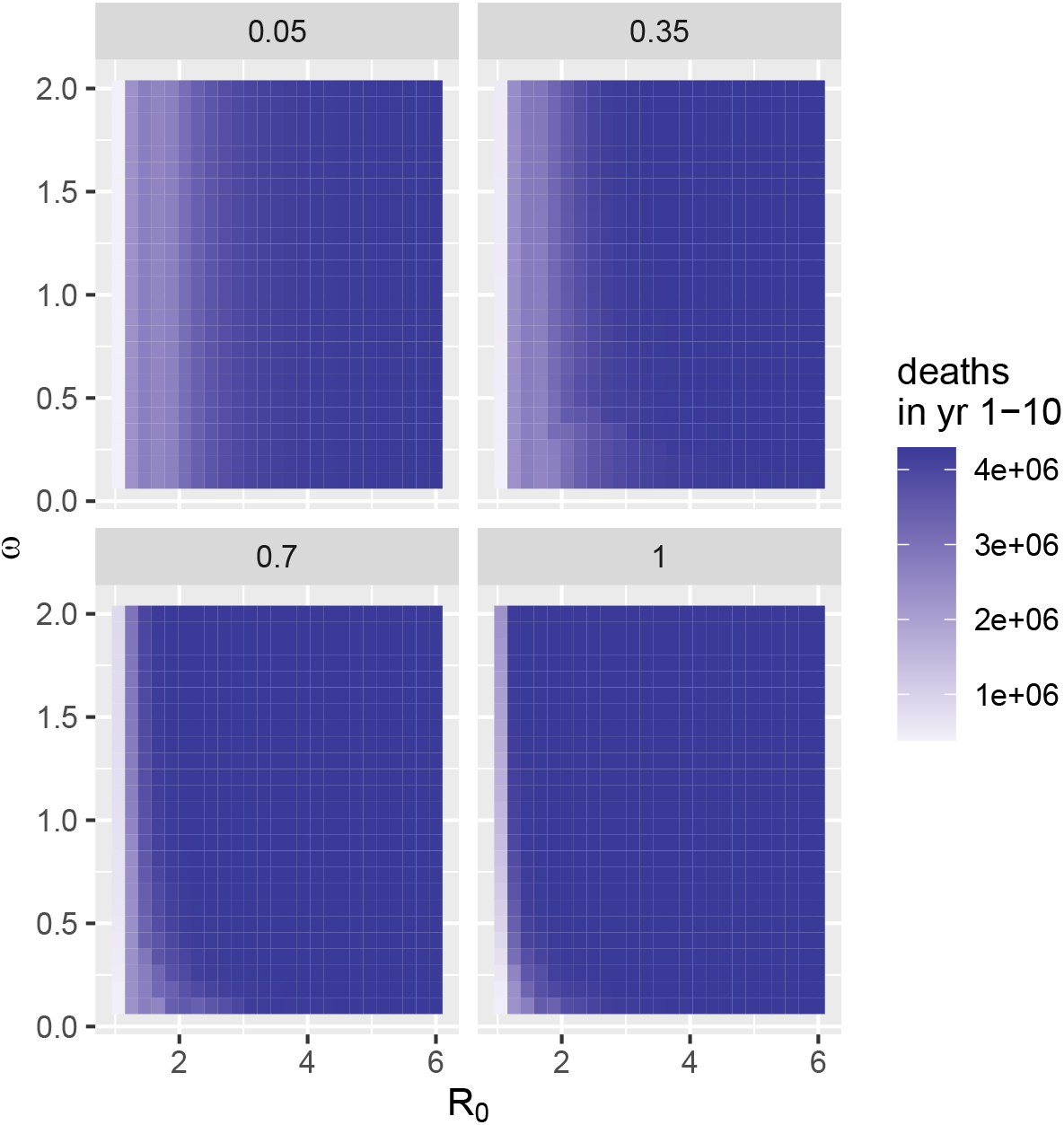
Number of infection-induced deaths in first ten years after emergence. When reinfection occurs quickly and is transmissible (high *ω* and *ρ*), the total number of infection-induced deaths is close to independent of *R*_0_.

### 3 Loss of immunity kernel

We estimate the duration of sterilizing immunity based on Callow et al. [5], in which six out of nine people who got infected at the start of the experiment were susceptible to reinfection one year later. We can calculate the mean of *ω* given this for exponentially distributed waning times setting the CDF equal to 2/3.

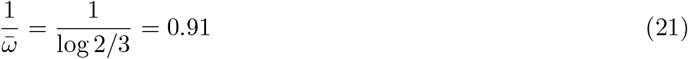

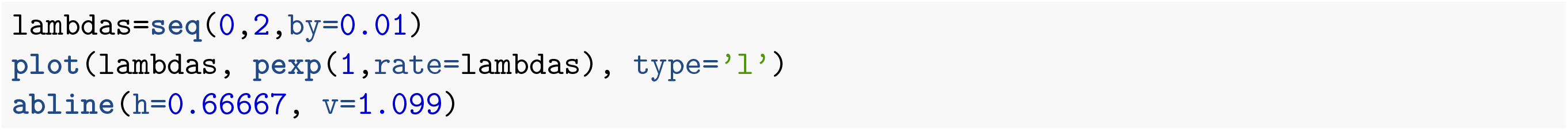

Given this, the estimate for *ω* is 1.099 and the mean waiting time is 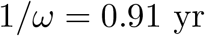.

We can also find the average duration of immunity for normally distributed waning times. We assume that the variance is the same as for the exponential model, 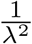 and the standard deviation is 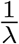.

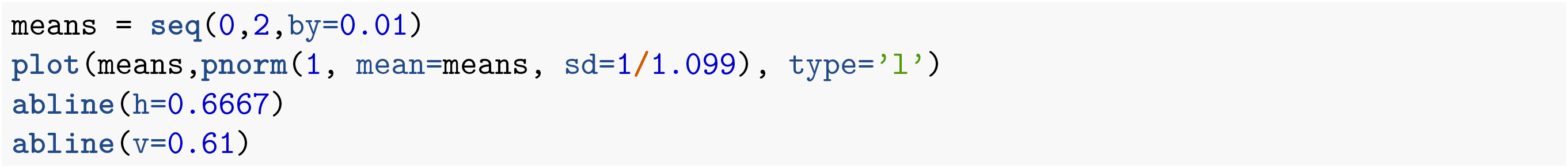

Here, we find the estimate for the mean of *ω* to be 0.61/yr, so the mean waiting time is 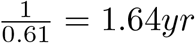.

### 4 Quarantine analysis

To better understand how waning of immunity impacts symptom-based quarantining in the first few years after disease emergence, we consider the following three phases. pexp(1, rate = lambdas)

**Figure 7:**
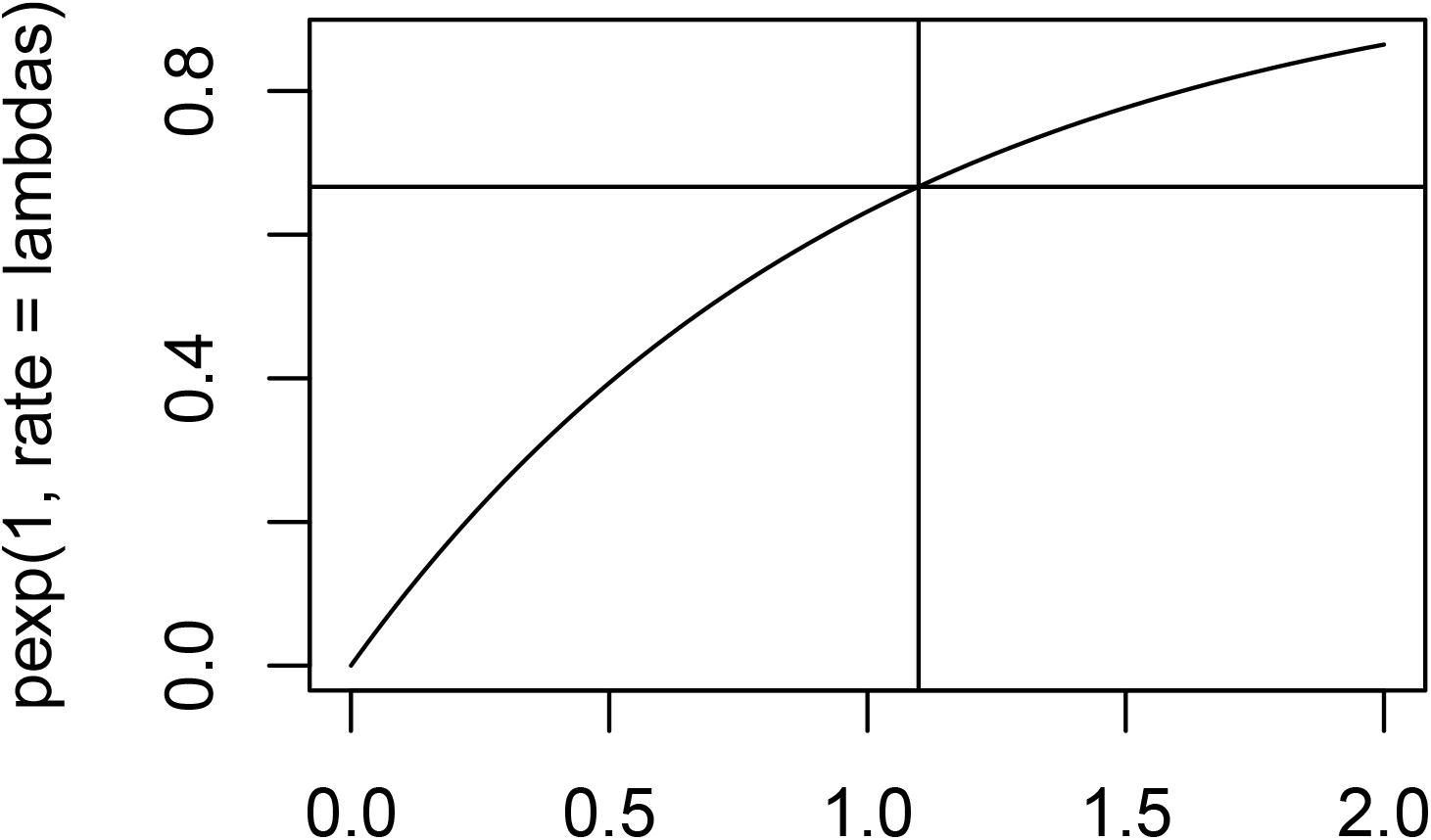
Estimated duration of immunity given exponentially distributed waning times

### 4.1 Phase 1

The initial epidemic, before control measures are introduced, burns through a proportion of the population, *p*_1_. At some point, let’s call it *t*_0_, we introduce highly effective quarantining, which breaks chains of transmission. Because the initial epidemic burns through the population quickly, we approximate the time of entry into *R*_1_ as being uniform. Additionally, since chains of transmission are broken upon the introduction of effective quarantining and contact tracing, we assume no new cases arise after *t*_0_ during Phase1. Therefore at *t*_0_ (or shortly after):

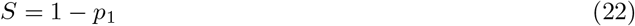

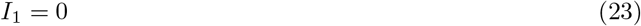

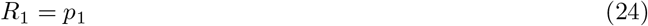

### 4.2 Phase 2

There is a time period during which the occasional immigrant case enters, but no chains of transmission start. Even if the immigrant with a primary or secondary infection infects a Susceptible person before being quarantined, that *I*_1_ is immediately quarantined and does not infect others. If an *R*_2_ person is infected, they are not quarantined but neither can they start a chain of transmission because any infection to S results in quarantine, and a transmission chain all among secondary (or later) cases will not happen because 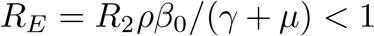. During this time period, people who have recovered from a primary infection are losing their immunity.

During this phase:

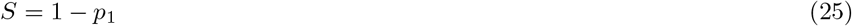

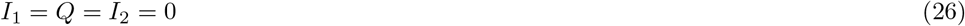

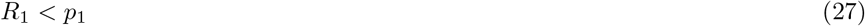

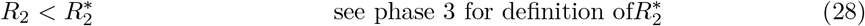

### 4.3 Phase 3

There is a threshold value of *R*_2_, let’s call it 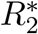, above which an immigrant case can lead to an outbreak of asymptomatic secondary infections, which acts as a reservoir for symptomatic primary infections. This occurs even in the presence of a strong quarantining program for symptomatic primary cases. If 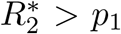, quarantining can continue working indefinitely. However, if 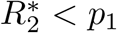, immune waning will lead to a sufficient build up of secondary susceptibles (in *R*_2_) to sustain an outbreak. In this scenario, the ‘safe time’ during which strong quarantining of primary cases can prevent outbreaks is influenced by: how fast immunity wanes (*ω*), how transmissible secondary cases are (*ρβ*_0_) and the proportion of the population that was infected in the initial outbreak (*p*_1_).

**Figure 8:**
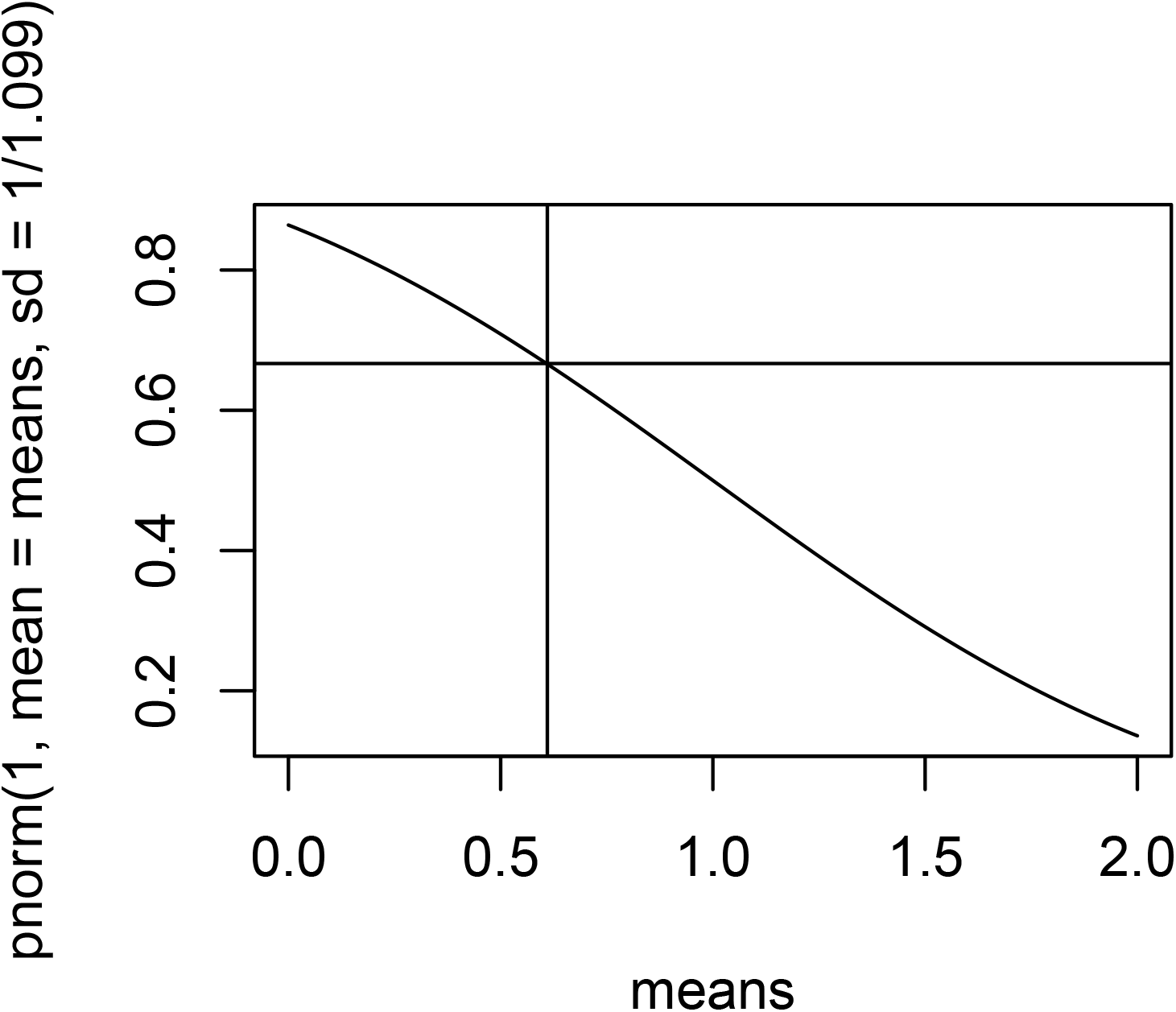
Estimated duration of immunity given normally distributed waning times

**Figure 9:**
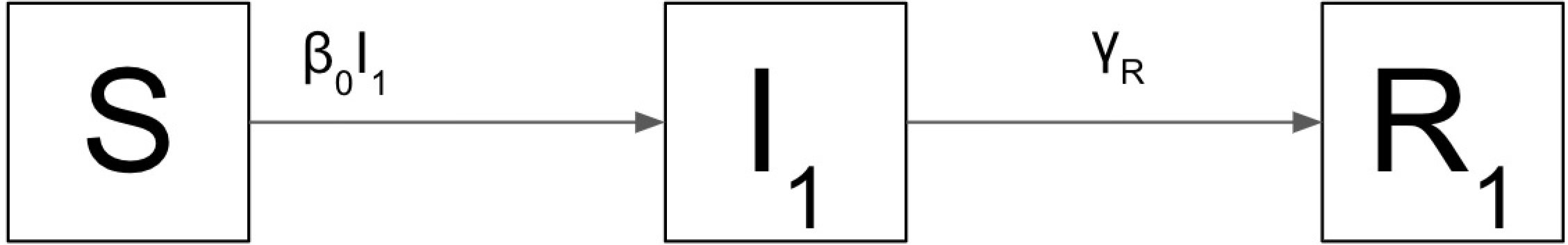
Phase 1 transmission

During this phase:

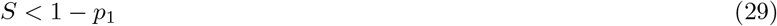

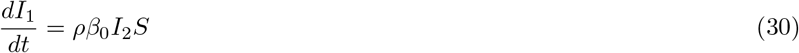

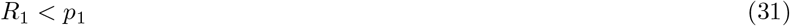

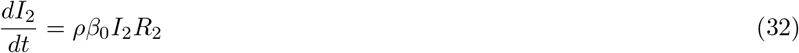

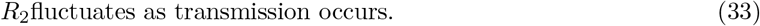

### 4.4 Window of safety

Here, we show results for the window of safety during which strict quarantining of primary cases will prevent an immigrant infection from leading to an outbreak for *R*_0_ = 6 and *R*_0_ = 1.5 to mimic a high but reasonable transmissionrate and a rate that may be more commensurate with the current scenario given social distancing.

**Figure 10:**
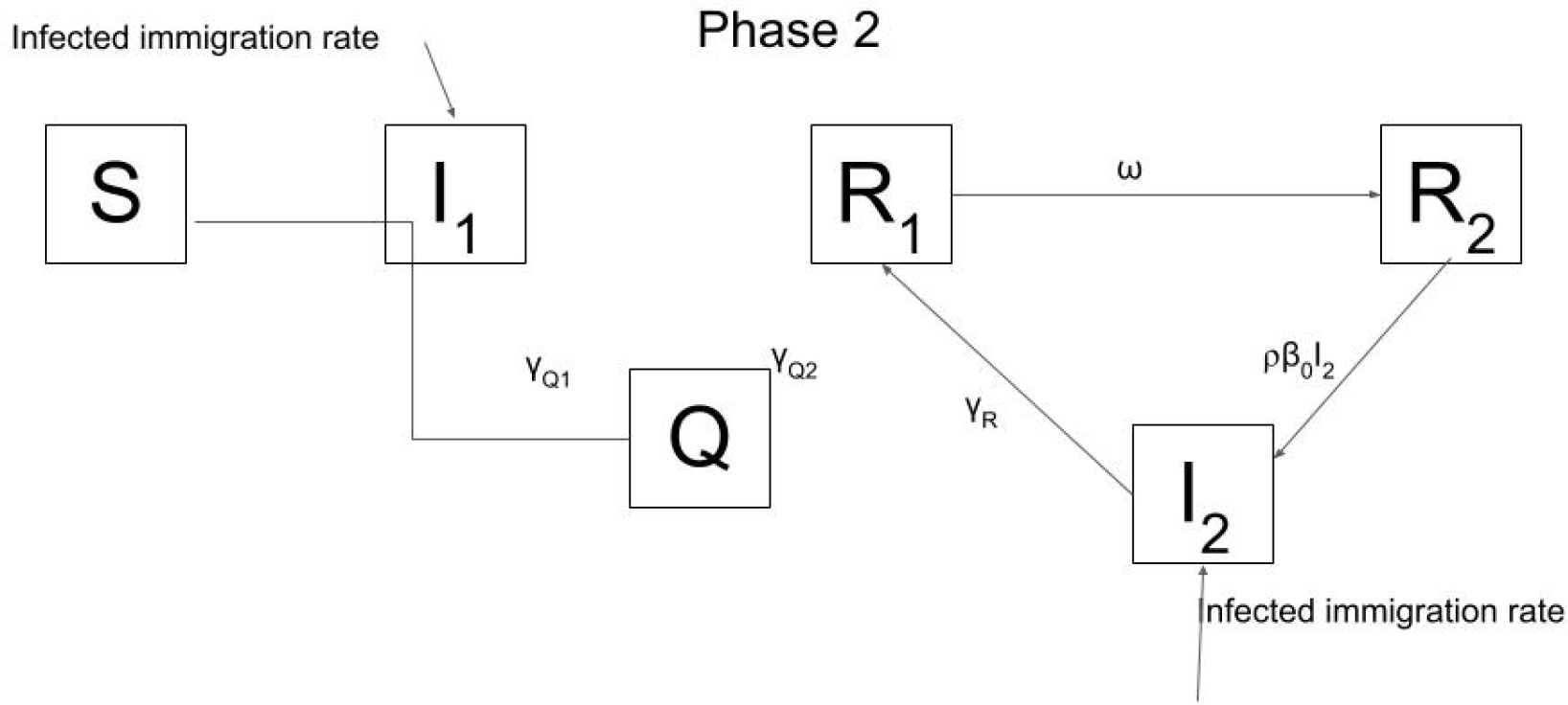
Phase 2 transmission

For *R*_0_ = 2 we see

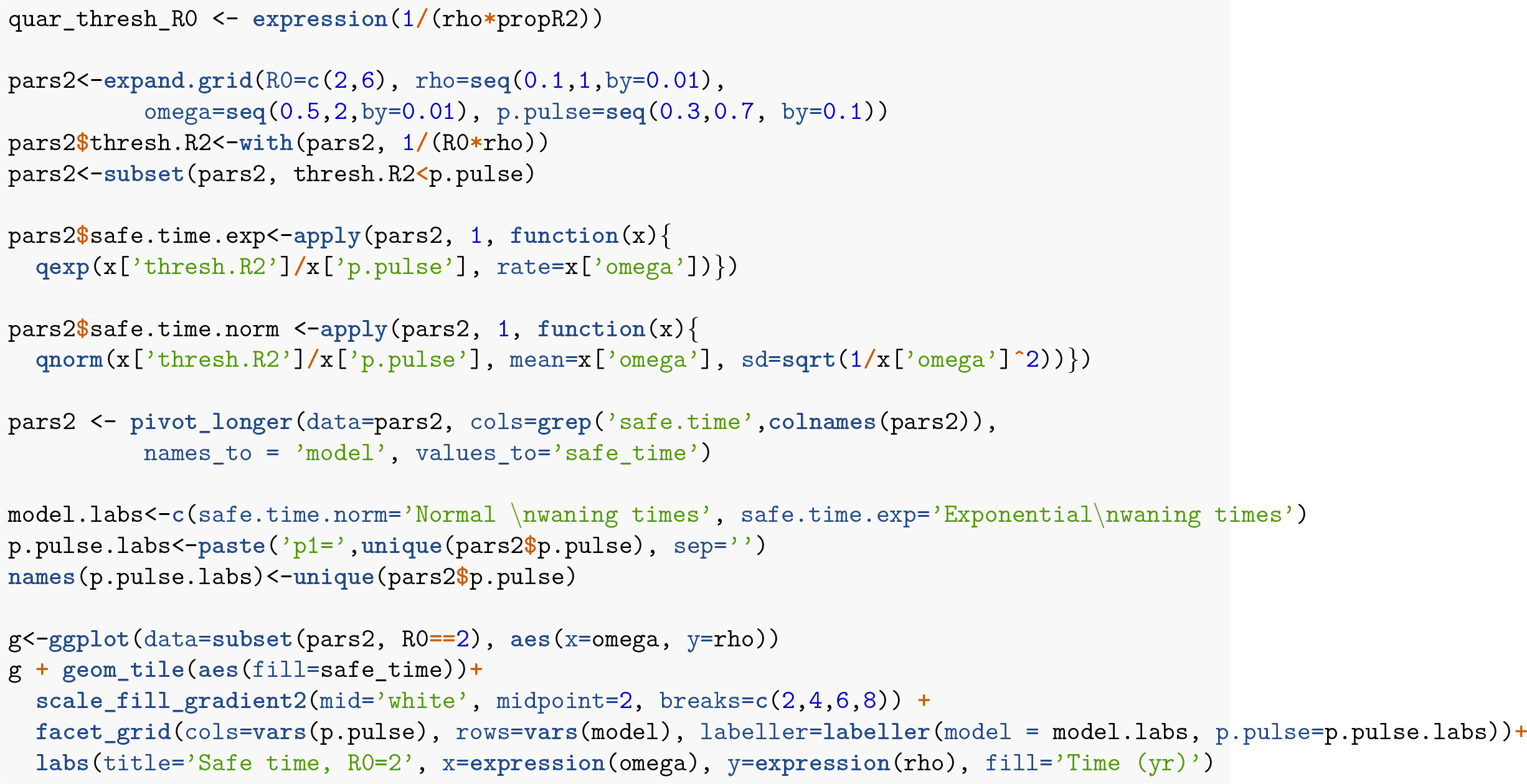

**Figure 11:**
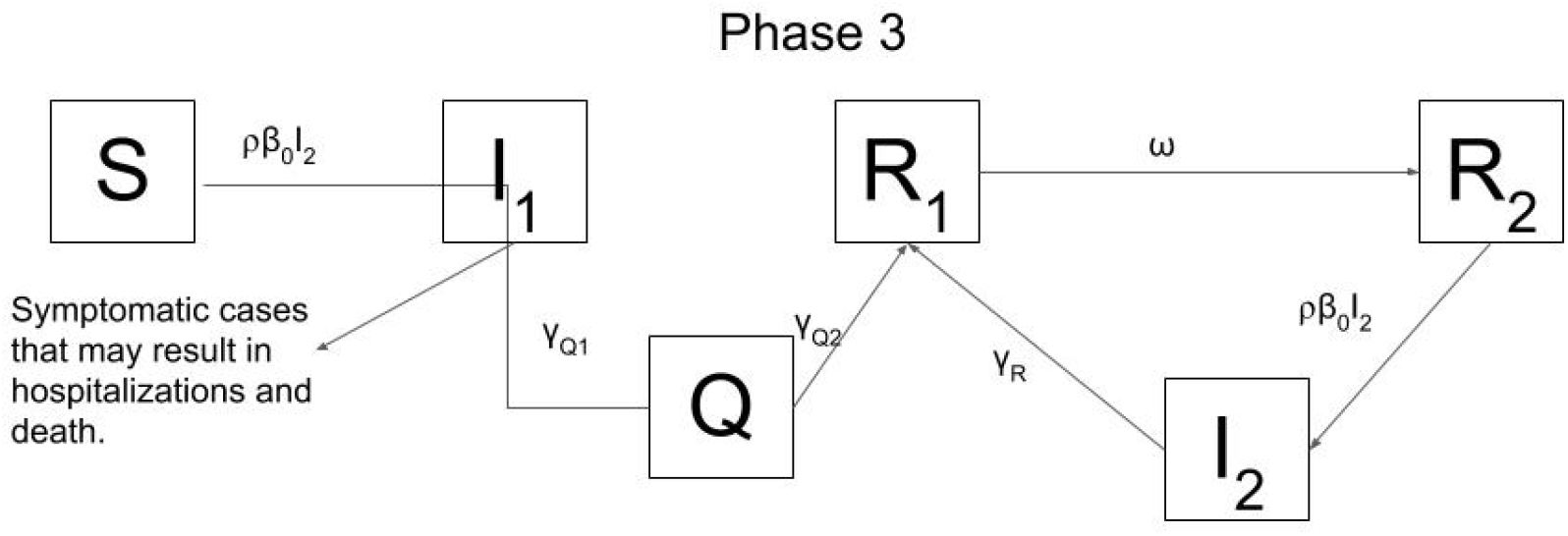
Phase 3 transmission

**Figure 12:**
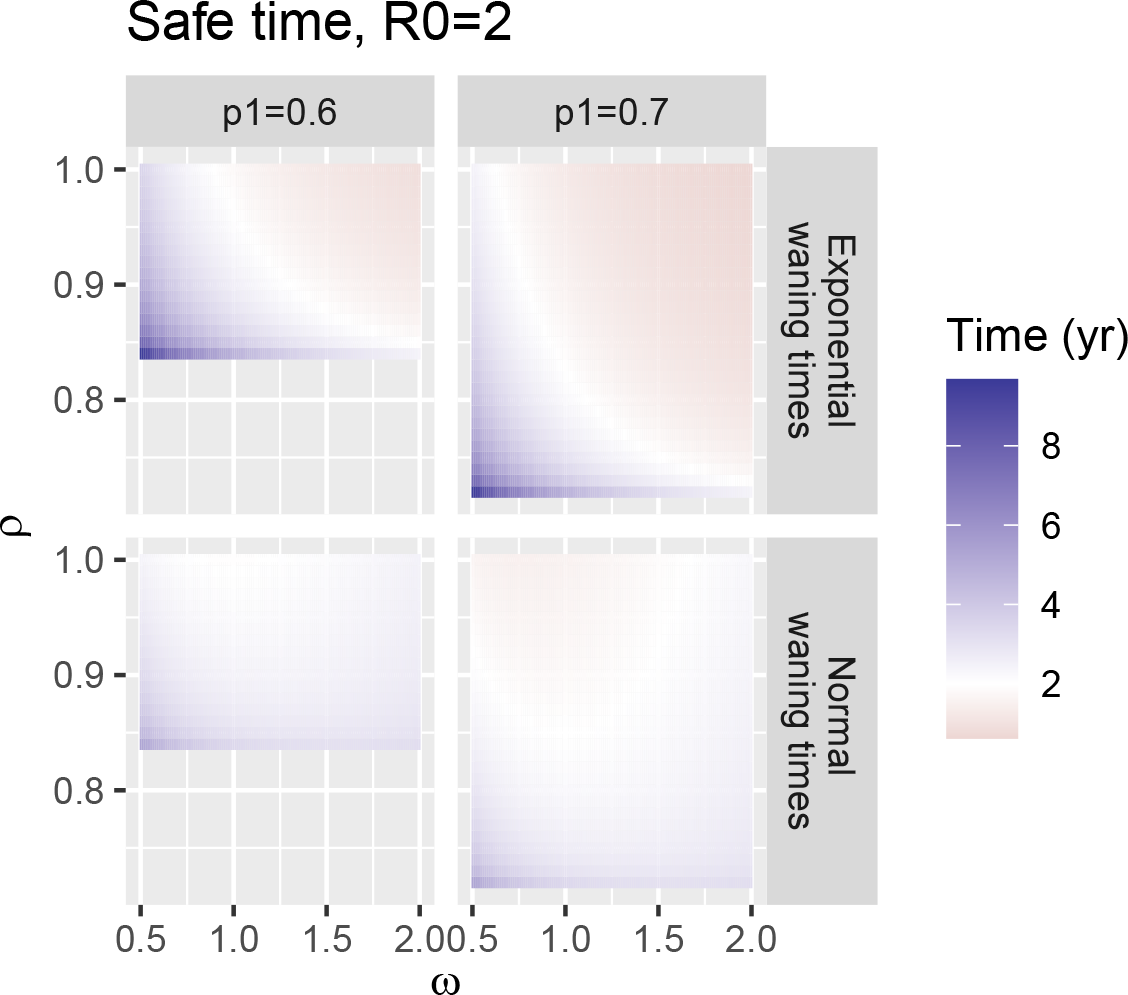
Safe time for *R*_0_ = 2

For *R*_0_ = 6, a smaller *p*_1_ can support an outbreak (as low as p1 = 0.3). The amount of time it takes for people’s immunity to wane sufficiently has a broader range.

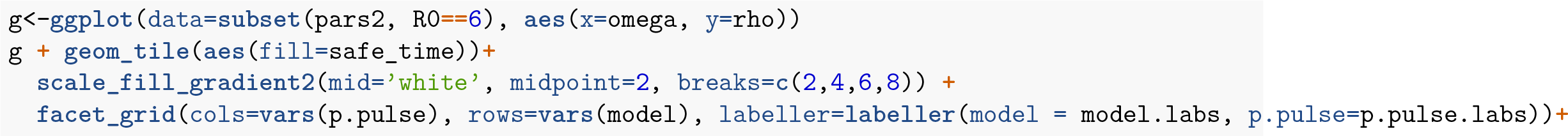
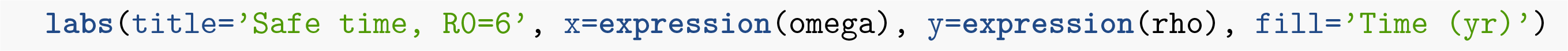

**Figure 13:**
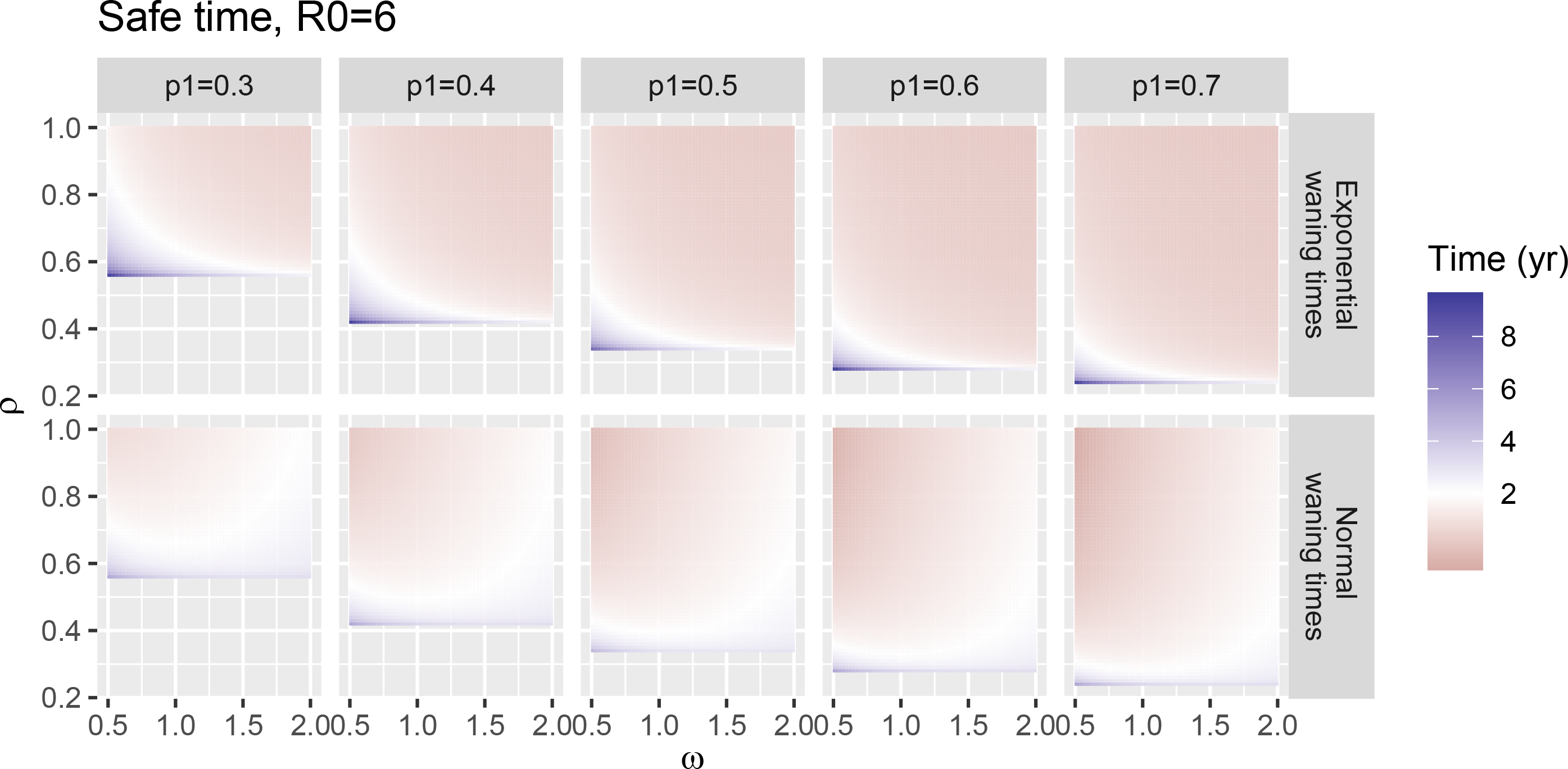
Safe time for *R*_0_ = 6

In reality, many primary cases have also been found to be asymptomatic, making symptom-based quarantine substantially more challenging and highlighting the need for asymptomatic surveillance to protect against transmission to vulnerable individuals [8].

## 5 Age-severity curves

We estimate the age-severity curves for the three HCoVs to have emerged in the past few decades using published data (CoV-2 [9], SARS CoV-1 [10], and MERS [11]). We then fit a generalized linear models to each data set to estimate a smoothed CFR as a function of age. We use a binomial model in which total cases in an age group is considered the number of trials, and the number of deaths is considered the number of ‘successes’. For MERS, we allow the function to be a third degree polynomial to account for the non-monotonicity of the data. We consider a second degree polynomial for SARS CoV-1 and −2, since the relationship between age and CFR appears monotonic in the data for these. Our code is included below.

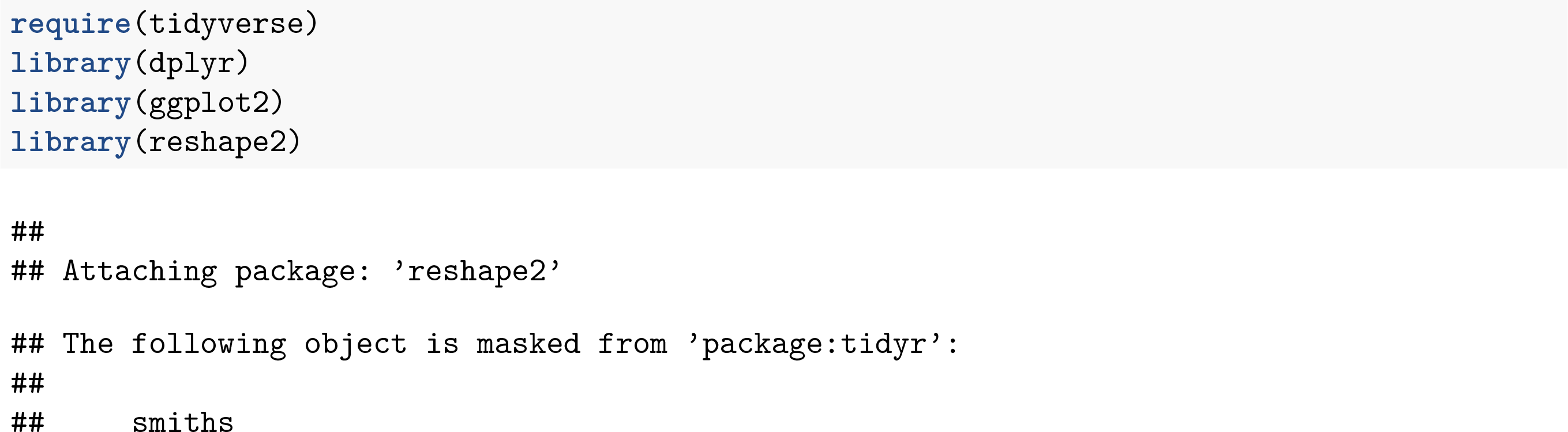

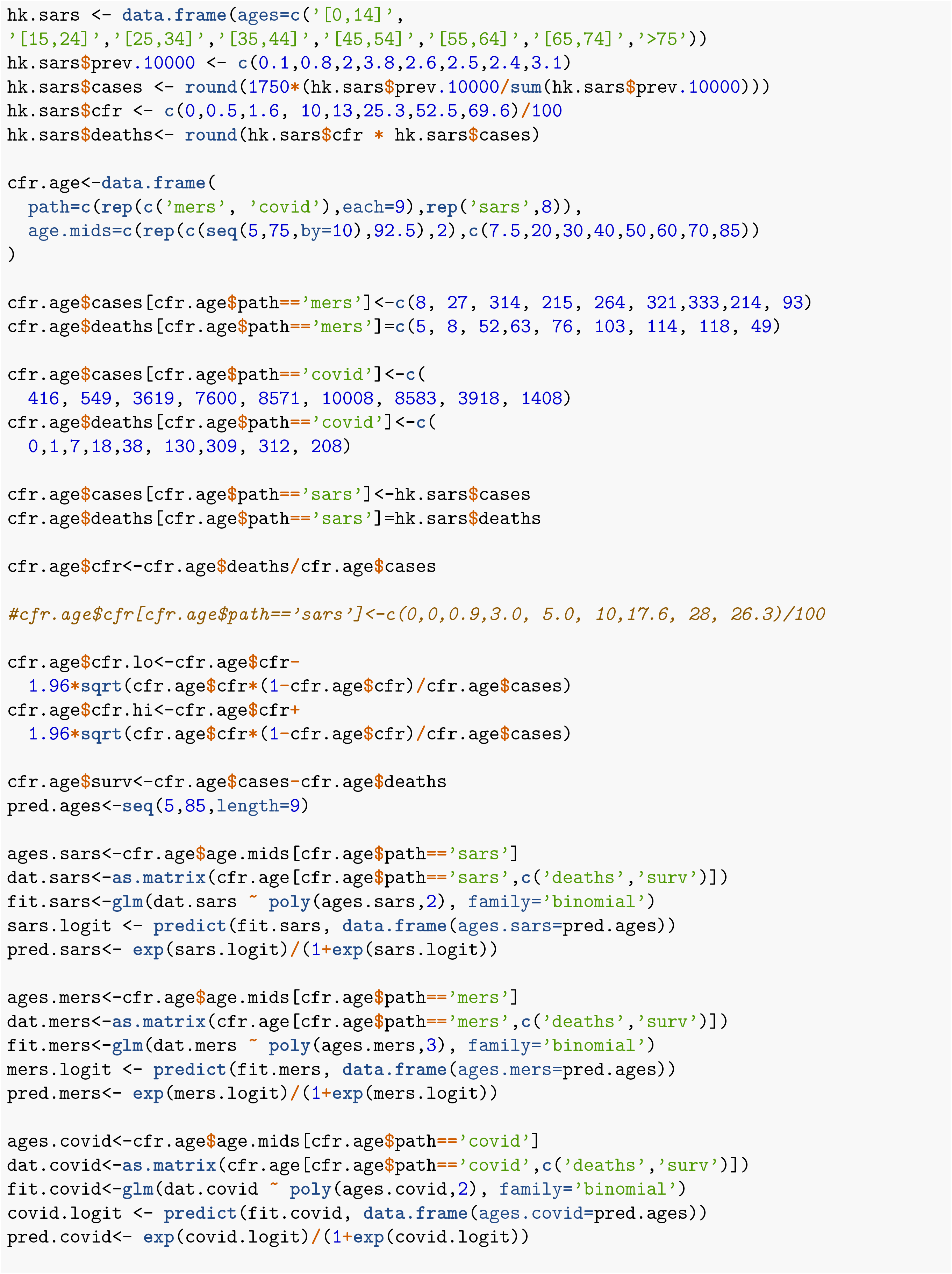

## 6 Code availability and reproducibility

All code will be available as an RMarkdown file.

## Notes

### Competing Interest Statement

The authors have declared no competing interest.

### Funding Statement

NIH

### Author Declarations

Analysis involves data from literature

